# High-Throughput Clinical Trial Emulation with Real World Data and Machine Learning: A Case Study of Drug Repurposing for Alzheimer’s Disease

**DOI:** 10.1101/2022.01.31.22270132

**Authors:** Chengxi Zang, Hao Zhang, Jie Xu, Hansi Zhang, Sajjad Fouladvand, Shreyas Havaldar, Feixiong Cheng, Kun Chen, Yong Chen, Benjamin S. Glicksberg, Jin Chen, Jiang Bian, Fei Wang

## Abstract

Clinical trial emulation, which is the process of mimicking targeted randomized controlled trials (RCT) with real-world data (RWD), has attracted growing attention and interest in recent years from the pharmaceutical industry. Different from RCTs which have stringent eligibility criteria for recruiting participants, RWD are more representative of real-world patients to whom the drugs will be prescribed. One technical challenge for trial emulation is how to conduct effective confounding control with complex RWD so that the treatment effects can be objectively derived. Recently many approaches, including deep learning algorithms, have been proposed for this goal, but there is still no systematic evaluation and practical guidance on them. In this paper, we emulate 430, 000 trials from two large-scale RWD warehouses, covering both electronic health records (EHR) and general claims, over 170 million patients spanning more than 10 years, aiming to identify new indications of approved drugs for Alzheimer’s disease (AD). We have investigated the behaviors of multiple different approaches including logistic regression and deep learning models, and propose a new model selection strategy that can significantly improve the performance of confounding balance of the participants in different arms of emulated trials. We demonstrate that regularized logistic regression-based propensity score (PS) model outperforms the deep learning-based PS model and others, which contradicts with our intuitions to a certain extent. Finally, we identified 8 drugs whose original indications are not AD (pantoprazole, gabapentin, acetaminophen, atorvastatin, albuterol, fluticasone, amoxicillin, and omeprazole), hold great potential of being beneficial to AD patients.

---

Pharmaceutical development of novel therapeutics for Alzheimer’s disease (AD) has consumed a large amount of resources over past decades but the majority of AD clinical trials have failed to produce positive results^1^. Drug repurposing, i.e., identifying novel indications for already approved drugs with well-defined safety and toxicity profiles, can potentially serve as a cost-effective way to accelerate AD drug development with a higher success rate^2^. Although repurposing drugs for AD has received increasing attention, no success has been reported on clinical sites^3^. One important reason is that existing efforts have been mostly based on pre-clinical (e.g., -omics, chemical, etc.) data, however, due to the complexity of the disease, these insights may not be directly translational to clinical settings.

On the other hand, large-scale real-world patient data (RWD), such as electronic health records (EHR) or administrative claims, has been accumulated in recent years and becoming readily available. Generating drug repurposing hypotheses from RWD through emulating randomized clinical trials (RCTs) with a causal analysis framework has demonstrated great potential in accelerating translation from bench to bedside for drug development and discovery^4–7^. This framework consists of two major steps: high-throughput RCT emulation for a large set of drug candidates using RWD, and estimation of the treatment effect of each drug candidate with causal analysis methods (such as the inverse probability of treatment re-weighting, or IPTW) for screening at scale. Due to the complexity of RWD, trial emulation with large-scale RWD has become a great touchstone for advanced AI algorithms, including machine learning or deep learning-based propensity score methods, for effective inference of treatment effects of drugs by adjusting for complicated confounding issues inherent within the observational data. As an example, recently an advanced deep learning-based long short-term memory with attention propensity score (PS) model^7^ showed superior performance in balancing covariates than logistic regression-based PS model when applied in the IPTW method. However, the superiority of these deep learning-based PS models still lacks systematic studies and validations on high-throughput trial emulations on different RWD databases, and whether it can be applied to AD is largely unknown.

In this study, with two large-scale RWD warehouses covering both electronic health records (EHR) and general claims, we systematically investigated the feasibility of generating AD repurposing hypotheses through high-throughput trial emulations, under an IPTW based framework with different ways of PS calculation. We emulated 430,000 RCTs for candidate drugs existing in RWD based on their impacts on the progression of patients with mild cognitive impairment (MCI) to AD. Inferring such treatment effects from large-scale RWD requires that different drug exposure groups to be balanced after IPTW with respect to high-dimensional baseline covariates^4,6–8^. Interestingly, we observed that the state-of-the-art deep learning-based PS model failed to balance the majority of our emulated trials. Specifically, these models usually result in insufficient overlap in different exposure groups, namely some patients in one exposure group have zero probability of being assigned to another group. This violates the basic positivity assumption in IPTW and can lead to balance failure with RWD. Related to this issue, we further demonstrate that building a PS model by purely optimizing the performance of predicting the likelihood of treatment can lead to failure in balancing covariates for patients in different treatment arms in trial emulations in our empirical studies. We, therefore, propose a new model selection strategy tailored for building machine learning models for PS calculation which yields significantly better balancing performance than existing practice. With the proposed strategy, we found that a simple regularized logistic regression-based PS model outperformed other complicated machine learning models including deep learning, and we are able to identify eight drugs including gabapentin, acetaminophen, atorvastatin, albuterol, fluticasone, pantoprazole, amoxicillin, omeprazle, with significant and consistent reduced risk of AD within 2 years, which can potentially serve as repurposing candidates for AD. Fig. 1 illustrates the overall pipeline of our proposed framework, which includes the following main steps.

**Figure 1.**
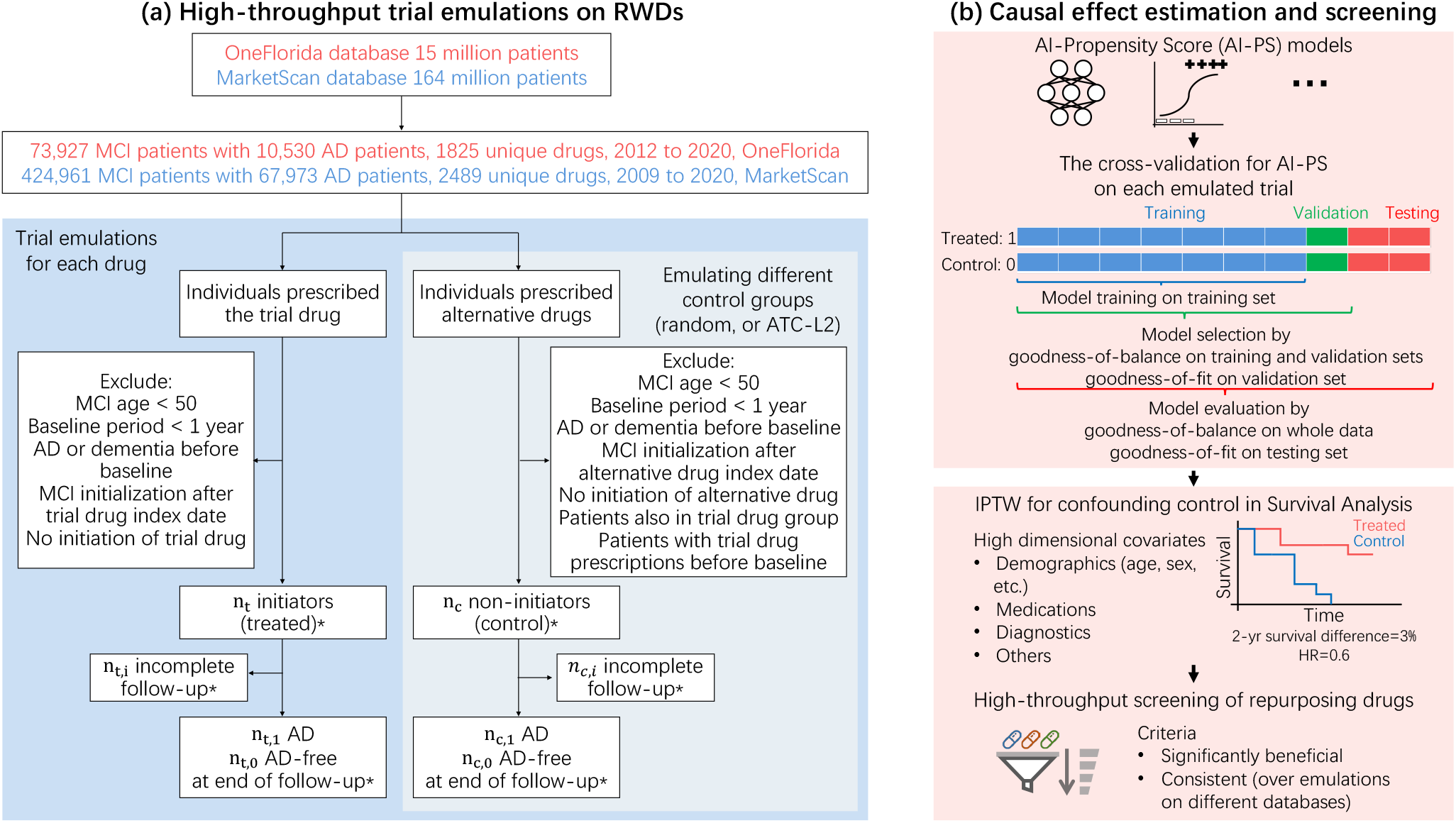
Overview of our high-throughput clinical trial emulation system for Alzheimer’s Disease drug repurposing driven by real-world data and machine learning. **(a)** High-throughput trial emulations of thousands of drug candidates were conducted on two large-scale and longitudinal real-world healthcare databases: OneFlorida and MarketScan. Target trial protocols (eligibility criteria, treatment strategies and assignment, follow-up, outcomes, etc.) were illustrated as a flow-chart (details in Method section). For each drug candidate, treated group consisted of patients who were prescribed with the trial drug, and control group was constructed by either random selection of alternative drug groups or using drug groups under the same second-level Anatomical Therapeutic Chemical classification codes (ATC-L2) as trial drug group. Hundreds of trials were emulated for each drug by constructing different control groups. *The number of patients in different groups and the outcomes were varied across emulated trials. MCI, mild cognitive impairment; AD, Alzheimer’s Disease. **(b)** Causal effect estimation for each emulated drug trial and high-throughput screening of drugs. State-of-the-art AI-based propensity score (AI-PS) models were used and compared. Novel cross-validation framework for AI-PS models was proposed for training, selecting, and evaluating AI-PS in terms of goodness-of-balance and goodness-of-fit performance. The optimally trained and selected AI-PS model is used for inverse probability of treatment re-weighting (IPTW) high-dimensional patient baseline covariates, including age, gender, disease comorbidities, medications, etc., for confounding control. AD event or censoring event were tracked within two-year follow-up period, and estimated treatment effects were quantified by adjusted two-year survival difference and adjusted hazard ratio (HR). Potentially repurposing drug candidates were selected if their estimated treatment effects were significantly beneficial and consistent over emulated trials on different databases.

First, we specify the protocols of targeted trials and their high-throughput emulations using two large-scale RWD warehouses, where we treat every single drug existing in RWD as a potential candidate (Fig. 1a). The details of the protocols are summarized in the Method section and their key components are provided in the extended data table 1. Briefly, the eligibility criteria of the treated groups of any emulated trial include MCI patients with age ≥ 50, with at least 1 year of records in the database before baseline (the date of the first prescription of the target drug) for collecting covariates, MCI diagnosis before baseline, and no AD or AD-related dementia diagnoses before baseline. For each target drug candidate, we emulate one hundred trials by constructing different control groups selected either from patients who took a random alternative drug or a similar drug under the same second-level Anatomical Therapeutic Chemical classification codes (ATC-L2) as the target drug. We further excluded patients from the control groups who were also in the treated group or took any trial drugs before baseline. All patients were followed up to 2 years or until AD diagnosis or loss to follow-up (censoring). There were over 4, 300 unique drugs (grouped by their major active ingredients) in the two databases we investigated and for each drug we emulated 100 trials, leading to 430, 000 (referred to as high throughput) emulated trials in total.

**Table 1.** Screening of drug candidates^a^, characteristics of high-throughput emulated trials, and their estimated treatment effects ^b^, OneFlorida, 2012-2020.

| Drug | Model | Balanced trials % | No. of treated | No. of control<sup>c</sup> | No. of unbalanced feat.<sup>d</sup> | No. of unbalanced feat. after IPTW | Adjusted 2-yr survival difference % (95% CI)<sup>a</sup> | Adjusted hazard ratio (95% CI)<sup>a</sup> |
| --- | --- | --- | --- | --- | --- | --- | --- | --- |
| Escitalopram | LR | 29 | 767 | 2301.0 | 45.2 | 3.1 | 3.7 (3.0,4.5)*** | 0.61 (0.55,0.67)*** |
|  | LSTM | 26 | 767 | 2301.0 | 22.7 | 3.4 | 5.0 (4.1,5.8)*** | 0.50 (0.45,0.57)*** |
| Mirtazapine | LR | 32 | 810 | 2430.0 | 53.3 | 2.8 | 2.4 (1.7,3.1)*** | 0.76 (0.68,0.86)*** |
|  | LSTM | 40 | 810 | 2430.0 | 24.7 | 2.6 | 3.4 (2.7,4.1)*** | 0.66 (0.59,0.73)*** |
| **Pantoprazole** | LR | 91 | 1100 | 2567.0 | 114.6 | 1.5 | 2.4 (2.2,2.6)*** | 0.57 (0.56,0.59)*** |
|  | LSTM | 93 | 1100 | 2502.1 | 38.1 | 2.0 | 2.6 (2.5,2.7)*** | 0.51 (0.50,0.53)*** |
| Meloxicam | LR | 23 | 675 | 2025.0 | 68.1 | 4.2 | 2.3 (2.2,2.4)*** | 0.53 (0.51,0.54)*** |
|  | LSTM | 0 | — | — | — | — | — | — |
| **Gabapentin** | LR | 59 | 1237 | 3260.3 | 89.6 | 1.5 | 2.1 (1.8,2.4)*** | 0.61 (0.58,0.64)*** |
|  | LSTM | 41 | 1237 | 2636.8 | 46.0 | 3.5 | 2.0 (1.6,2.4)*** | 0.62 (0.59,0.66)*** |
| Sertraline | LR | 28 | 709 | 2127.0 | 48.5 | 3.1 | 2.1 (1.3,2.8)*** | 0.83 (0.73,0.92)** |
|  | LSTM | 33 | 709 | 2127.0 | 27.3 | 3.4 | 2.9 (2.1,3.6)*** | 0.75 (0.68,0.82)*** |
| Trazodone | LR | 66 | 1126 | 3370.9 | 95.9 | 2.3 | 1.6 (0.9,2.2)** | 0.92 (0.82,1.01)<sup>ns</sup> |
|  | LSTM | 55 | 1126 | 3378.0 | 38.8 | 2.7 | 2.7 (2.1,3.3)*** | 0.74 (0.67,0.82)*** |
| **Acetaminophen** | LR | 57 | 1837 | 4864.9 | 45.0 | 1.0 | 1.4 (1.2,1.7)*** | 0.76 (0.73,0.78)*** |
|  | LSTM | 84 | 1837 | 3568.0 | 45.9 | 1.7 | 1.4 (1.3,1.6)*** | 0.76 (0.74,0.77)*** |
| **Atorvastatin** | LR | 96 | 1674 | 3151.6 | 67.5 | 1.0 | 1.2 (1.1,1.4)*** | 0.79 (0.77,0.81)*** |
|  | LSTM | 68 | 1674 | 2700.1 | 25.9 | 1.8 | 1.7 (1.6,1.9)*** | 0.71 (0.69,0.72)*** |
| **Albuterol** | LR | 22 | 1045 | 2126.6 | 85.3 | 3.8 | 1.2 (0.9,1.5)*** | 0.78 (0.74,0.83)*** |
|  | LSTM | 26 | 1045 | 2282.1 | 55.3 | 3.9 | 1.0 (0.7,1.3)*** | 0.80 (0.75,0.86)*** |
| Lisinopril | LR | 47 | 950 | 1864.4 | 46.4 | 3.0 | 0.8 (0.7,1.0)*** | 0.82 (0.80,0.84)*** |
|  | LSTM | 29 | 950 | 1883.2 | 25.7 | 3.8 | 0.6 (0.4,0.9)*** | 0.85 (0.81,0.89)*** |
| **Fluticasone** | LR | 41 | 903 | 2709.0 | 82.6 | 3.9 | 0.7 (0.4,1.0)*** | 0.90 (0.84,0.94)*** |
|  | LSTM | 0 | — | — | — | — | — | — |
| **Amoxicillin** | LR | 13 | 668 | 2004.0 | 57.9 | 3.8 | 0.6 (0.3,0.9)* | 0.88 (0.82,0.95)* |
|  | LSTM | 0 | — | — | — | — | — | — |
| **Omeprazole** | LR | 48 | 917 | 2147.9 | 88.0 | 3.1 | 0.5 (0.4,0.7)*** | 0.88 (0.85,0.91)*** |
|  | LSTM | 0 | — | — | — | — | — | — |
| Famotidine | LR | 77 | 842 | 2450.7 | 93.5 | 2.6 | 0.1 (-0.1,0.2)<sup>ns</sup> | 0.99 (0.95,1.02)<sup>ns</sup> |
|  | LSTM | 24 | 842 | 2457.8 | 25.9 | 4.0 | 0.0 (-0.3,0.2)<sup>ns</sup> | 1.00 (0.96,1.05)<sup>ns</sup> |
| Folic acid | LR | 11 | 844 | 2532.0 | 60.8 | 3.0 | -0.3 (-0.7,0.3)<sup>ns</sup> | 1.06 (0.95,1.16)<sup>ns</sup> |
|  | LSTM | 0 | — | — | — | — | — | — |
| Losartan | LR | 28 | 801 | 2193.0 | 73.0 | 3.5 | -0.3 (-0.4,-0.2)*** | 1.00 (0.98,1.01)<sup>ns</sup> |
|  | LSTM | 25 | 801 | 2209.8 | 40.1 | 2.8 | 0.0 (-0.1,0.1)<sup>ns</sup> | 0.93 (0.91,0.95)*** |
| Metoprolol | LR | 19 | 892 | 2466.8 | 47.8 | 3.7 | -0.6 (-0.9,-0.4)*** | 1.18 (1.10,1.29)*** |
|  | LSTM | 0 | — | — | — | — | — | — |
| Ergocalciferol | LR | 13 | 996 | 2988.0 | 92.3 | 3.8 | -0.6 (-1.0,-0.2)<sup>ns</sup> | 1.14 (1.05,1.23)* |
|  | LSTM | 0 | — | — | — | — | — | — |
| Amlodipine | LR | 44 | 930 | 2789.6 | 51.8 | 3.0 | -0.7 (-0.8,-0.6)*** | 1.18 (1.15,1.22)*** |
|  | LSTM | 28 | 930 | 2790.0 | 32.9 | 3.9 | -0.6 (-0.8,-0.4)*** | 1.17 (1.12,1.23)*** |
| Aspirin | LR | 87 | 1532 | 4573.5 | 50.3 | 1.7 | -1.4 (-1.6,-1.2)*** | 1.31 (1.27,1.36)*** |
|  | LSTM | 78 | 1532 | 4570.9 | 48.6 | 2.6 | -1.6 (-1.8,-1.4)*** | 1.33 (1.28,1.37)*** |

Next, we propose a new model selection strategy tailored for training, selecting, and evaluating PS calculation methods, by taking into account both goodness-of-balance and goodness-of-fit (Fig. 1b). In particular, we first randomly partition each emulated trial into mutually exclusive training, validation, and testing sets, and then a) train the PS model on the training set, b) select the best model according to the goodness-of-balance measure on the training and validation combined sets, and the goodness-of-fit measure on the validation set, and c) evaluate the selected best PS model according to the goodness-of-balance measure on the whole dataset and the goodness-of-fit measure on the test set. We quantify the goodness-of-balance by the standardized mean difference (SMD) and the goodness-of-fit by the area under the receiver operating characteristic (AUC). We tested 4 different PS calculation models including regularized logistic regression, long short-term memory network (LSTM, with attention mechanisms)^7^, gradient boosted decision trees (GBDT)^9–11^, and multi-layer perceptrons^12,13^, and observed that i) all these models learned and selected by our proposed strategy balanced more emulated trials than existing model selection strategies, and ii) with our strategy, complicated machine learning models such as LSTM and GBDT did not necessarily outperform the simple regularized logistic regression PS model.

Finally, we estimate the treatment effects from high-throughput emulated trials and based on which we screen and prioritize potential candidates of non-AD drugs that can be repurposed for treating AD (Fig. 1b). We compute the stabilized IPTW weights by the aforementioned PS models and use these learned weights to re-weight each emulated trial. We compute the number of unbalanced covariates of emulated trials by SMD before and after IPTW. We estimate the treatment effects (adjusted 2-year survival difference and adjusted hazard ratio) of successfully balanced trials after IPTW. We propose two criteria, significant benefits, and consistency, to screen and prioritize drug candidates, based on which eight drugs were identified which show significantly beneficial effects on AD and their estimated beneficial effects are consistent over a large number of balanced emulated trials in both databases.

## Results

### Our model selection for propensity score calculation results in better balancing

We construct treatment groups consisting of eligible patients (Methods) for each unique drug ingredient existing in our databases and emulate trials for all of them. For each emulated drug trial, its treatment arm consists of patients from trial drug group, and its control arm is composed by either patients randomly selected from drug groups other than trial group, or patients from drug groups wherein drugs are under the same second-level Anatomical Therapeutic Chemical classification codes (ATC-L2) as the trial drug. To achieve statistical significance, we emulated 100 trials for each drug trial consisting of 50 emulations by constructing random control groups and 50 emulations by constructing ATC-L2 control groups. Taking the OneFlorida database (see Data Section) as our discovery set, we included 73, 927 patients with MCI diagnosis from 2012 to 2020 (Fig. 1a). We found 1, 825 unique drug ingredients and emulated 182, 500 trials. We finally targeted at 66 drugs with 6, 600 emulated trials of which each treatment group has ≥ 500 patients. For each emulated trial, we randomly partitioned the data into mutually exclusive training, validation and testing subsets as standard practice. All PS calculation models were trained on the same training set, and the best-estimated model was selected by following three strategies: (a) goodness-of-fit performance on the (out-of-sample) validation set, quantified by the area under the receiver operating characteristic curve (AUC) score^8,14–16^; (b) goodness-of-balance performance on the validation set, quantified by the maximum value of SMD scores over all baseline covariates after IPTW^7^; and (c) our proposed strategy, which leverages goodness-of-balance on the training and validation combined set, and goodness-of-fit on the validation set (Method Algorithm 1). We evaluated the performance of selected models from two aspects: (i) the goodness-of-balance, which measures how similar the baseline covariates of different exposure groups are after IPTW on the whole data, and (ii) the goodness-of-fit, which measures how good the learned PS model predicts on the unseen test data (See Method Algorithm 2). Of note, the goodness-of-balance is the single most important criterion for evaluating trial emulations. We considered one covariate as balanced if its SMD value ≤ 0.1^17^, and one emulated trial before/after IPTW is balanced if the ratio of unbalanced features among all covariates before/after IPTW ≤ 2%^7^. The goodness-of-fit was evaluated by the AUC score on the (unseen) test data, which is the most commonly used model evaluation strategy for machine learning models in general for binary classification.

Figure 2 shows the proportion of successfully balanced trials (≥ 10% among all emulations) for different drugs after IPTW by different PS models including: a) regularized logistic regression-based PS models (LR-PS), b) deep learning based-PS model using long short-term memory network with attention mechanisms (LSTM-PS)^7^, which were trained and selected by different model selection strategies as described above. Please refer to Extended Data Fig. 1 for the other PS models, including multi-layer perceptron based PS model (MLP-PS)^12,13^, and gradient boosted tree-based PS model (GBT-PS)^9–11^. We observed that PS models built with our proposed model selection strategy outperformed models selected by other strategies in terms of goodness-of-balance. The previous model selection strategies, either according to AUC (yellow bars in Fig. 2) or SMD (blue bars in Fig. 2) on validation data, failed to balance a large proportion of emulated trials (≤ 50%). By contrast, our strategy (red bars in Fig. 2) balanced many more emulated trials by large margins on all trial drugs. Taking atorvastatin as an example (Fig. 2a), LR-PS selected by our strategy balanced 96% [95% confidence interval (CI) 92% − 99%] of all emulated trials, which was much better than the AUC-based strategy (39%, 95% CI 30% −48%) and the SMD-based strategy (47%, 95% CI 37% −57%). The same phenomenon was observed on atorvastatin trials balanced by LSTM-PS (Fig. 2b), where our strategy (68%, 95% CI 58% −77%) balanced many more trials than AUC-(22%, 95% CI 14% −31%) or SMD-based strategy (53%, 95% CI 43% −63%). In addition, we also compared the performance of PS models built with different model selection strategies in terms of goodness-of-fit measured by AUC score on unseen test data. We observed that all of the above PS models selected by our proposed strategy achieved test AUC on par with models selected by AUC-on-validation strategy, and on par with or better than models selected by SMD-on-validation strategy (Extended Data Figs. 2 and 3). In summary, PS models selected by our proposed strategy balanced many more emulated trials than existing practice, and at the same time showed good generalized prediction performance on unseen data, and the proposed model training, selection, and testing processes were summarized in Algorithm 1 and 2.

**Figure 2.**
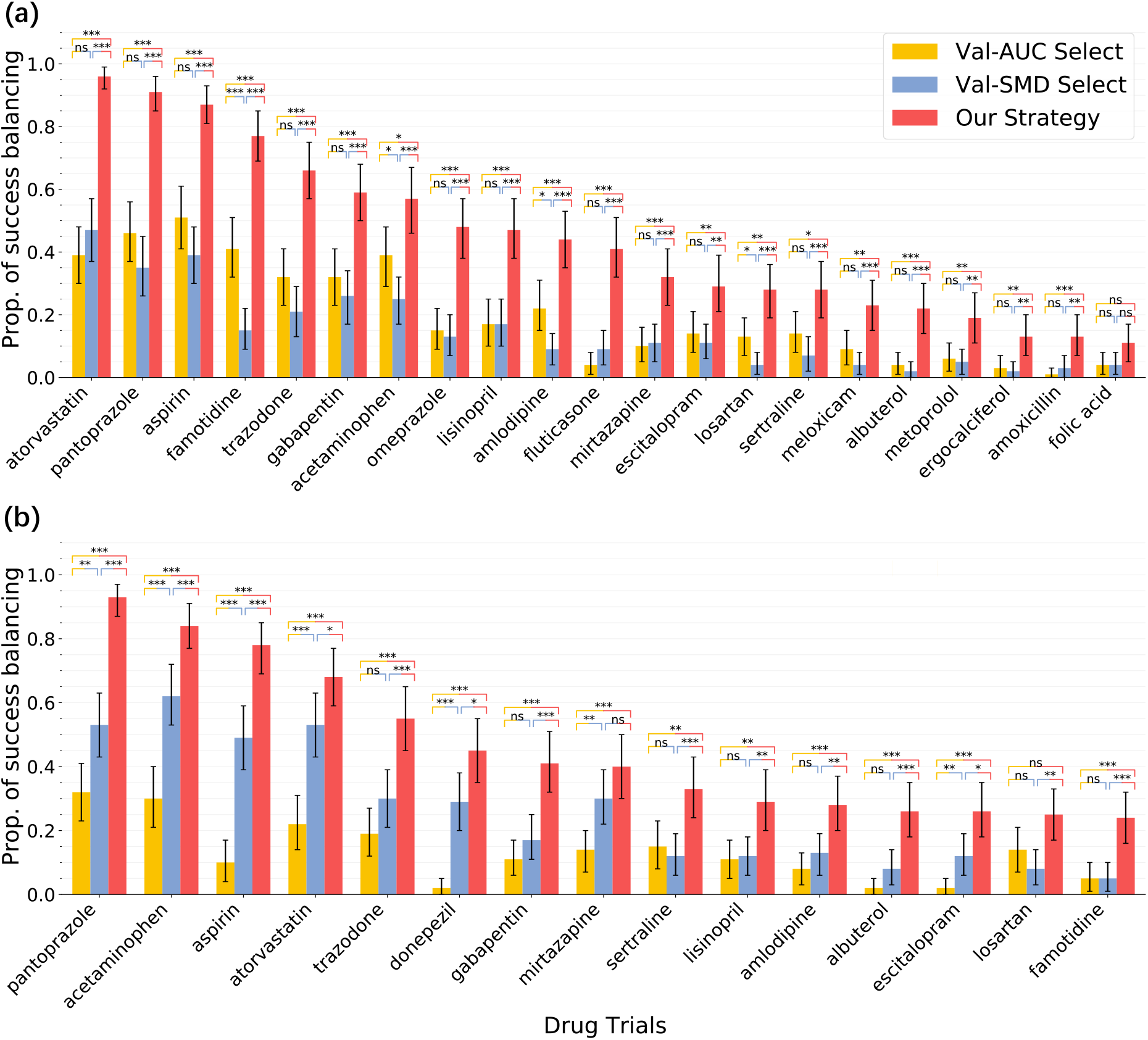
The proportion of successfully balanced drug trials, OneFlorida database, 2012-2020. PS models (a) LR-PS and (b) LSTM-PS selected by our model selection strategy balanced many more emulated trails than existing practice. Different color bars from left to right denote balancing performance by the best PS model selected under different strategies: AUC score on the validation set, maximum SMD after IPTW on the validation set, and our model selection strategy based on both the number of unbalanced covariates after IPTW on the training and validation combined set and AUC score on the validation set. We reported drugs with ≥ 10% balanced trials. The error bars indicate 95% confidence intervals by 1000-times bootstrapping. The (two-sided) independent two-samples T-test for testing the means of each two bars, and *, *p <* 0.05; **, *p <* 0.01; ***, *p <* 0.001; not significant (ns), *p ≥* 0.05; LR-PS, regularized logistic regression-based propensity score models; LSTM-PS, long short-term memory network with attention mechanisms-based propensity score models^7^; AUC, area under the receiver operating characteristic curve; SMD, standardized mean difference; IPTW, inverse probability of treatment re-weighting.

**Figure 3.**
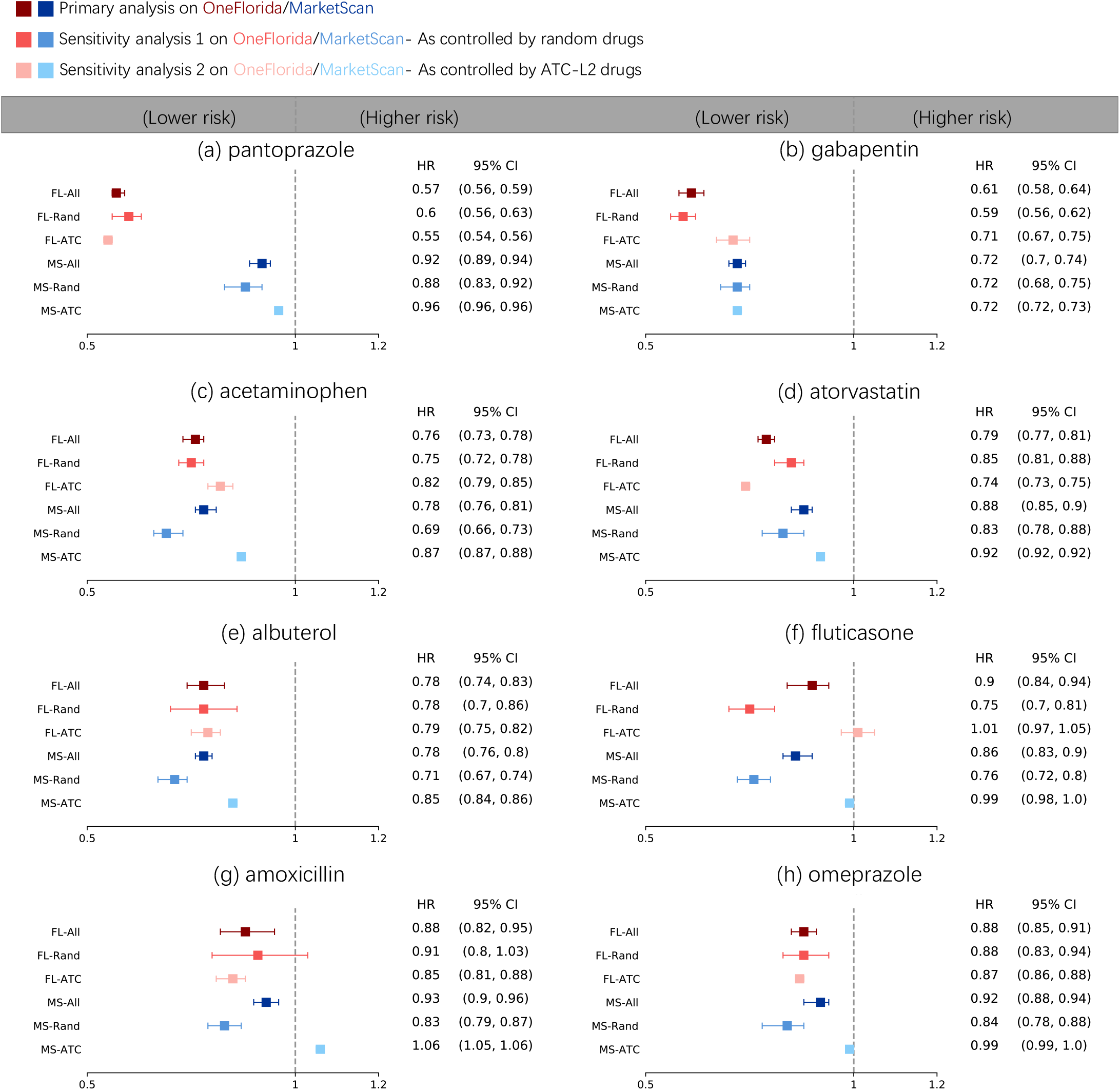
Eight repurposable drug candidates for AD with adjusted hazard ratios and 95% confidence intervals. Trial emulations of these eight drugs (a-h) were performed using OneFlorida (FL) and MarketScan (MS) data separately. For each drug, treated groups consisted of patients who prescribed the trial drug (eligibility criterion in the Methods section), and control groups were built by either: (1) randomly selecting alternative drug groups, or (2) using drug groups under the same second-level Anatomical Therapeutic Chemical classification codes (ATC-L2) as the trial drug. The primary analysis emulated 100 trials consisting of 50 random control groups and 50 ATC-L2 control groups (FL-All and MS-All), and two sensitivity analyses were using only random controls (FL-Rand and MS-Rand) or only ATC-L2 controls (FL-ATC and MS-ATC). The best regularized logistic regression-based propensity score (LR-PS) model selected by our proposed model selection strategy was used to adjust for 267-dimensional baseline covariates for each emulation. Mean hazard ratio (HR) of balanced emulated trials with 1,000-bootstrapped 95% confidence interval were reported.

### Does deep learning based models perform better?

Recently deep learning-based models have demonstrated great promises in various applications and researchers have proposed to apply these models for PS calculation in trial emulation^7^. We evaluated the performance of the PS calculation model based on the long short-term memory network with attention mechanisms (LSTM-PS) used in Liu et al.^7^ on our data, and observed that LSTM-PS did not necessarily outperform simple LR-PS. As shown in Fig. 2, the LR-PS model selected by our model selection strategy balanced 21 drugs of which ≥ 10% emulated trials were successfully balanced, while the LSTM-PS model only balanced 15 drugs. The LR-PS also identified more drug candidates than MLP-PS (9 drugs) and GBT-PS (9 drugs) as illustrated in Extended Data Fig. 1. Furthermore, we compared the balancing performance of LR-PS versus LSTM-PS by comparing the number of unbalanced features before and after IPTW (the 6th and 7th columns in Table 1), from which we observed that both LR-PS and LSTM-PS can greatly reduce the number of unbalanced features after re-weighting. However, the number of unbalanced features after IPTW by LSTM-PS similar to or even worse than the LR-PS model (LSTM rows v.s. LR rows in the 7th column). Moreover, LSTM-PS introduced additional biases by consistently under-estimating the number of unbalanced features even before re-weighting (LSTM rows v.s. LR rows in the 6th column in Table 1), which could be originated from the fact that LSTM-PS compressed the original covariate space through the learned attention weights, and the SMD scores were evaluated on the compressed covariates.

To test the generalizability of our conclusion, we further validated our proposed strategy on the MarketScan data, which is a national healthcare insurance claims database (see Data Section). Following the same procedures as we did with the OneFlorida data, we identified a total of 424, 961 MCI patients from 2009 to 2020 and among which, there were 2, 489 unique drug ingredients. We emulated 24, 600 trials for 246 drugs which had ≥ 500 patients in their respective treated groups. With the MarketScan data, we were able to obtain the same conclusions: (a) our model selection strategy built better PS models which balanced more trials than existing model selection strategies over different PS classes; (b) with our proposed strategy, conventional models such as LR-PS outperforms deep learning models such as LSTM-PS^7^ (Extended Data Fig. 4). In the following, we applied our strategy into the process of identification of repurposing drug candidates for AD via high-throughput trial emulation.

### High-throughput screening of repurposing drug candidates for AD

With our proposed model selection strategy and the LR-PS model, we have emulated 430,000 drug trials on two large-scale RWD warehouses (Fig. 1a). The adjusted 2-year survival difference and adjusted hazard ratio (HR) with AD onset as the outcome event obtained from these emulated trials are demonstrated in Fig. 1b. The repurposable drug candidates were identified according to the following two criteria: (i) beneficial effects, meaning the estimated treatment effect of balanced trials for any target drug should be significantly beneficial for MCI to AD progression (≥ 10% emulations were balanced after IPTW, the sample mean of adjusted 2-yr survival difference *>* 0, adjusted HR *<* 1, and P-value *<* 0.05); and (ii) consistency, the estimated treatment effects of each drug candidate from both RWD warehouses (both EHRs and administrative claims) should be beneficial for MCI to AD progression. Table 1 summarizes the screening process and emulated trials: with the first criterion we were able to identify 14 drugs from the OneFlorida data (Table 1, marked in shaded color) and 28 drugs from the Marketscan data (Supplementary Table 3), among which 8 drugs showed consistent beneficial effects on both data sets (Table 1, marked in bold).

We highlight these eight identified repurposable drug candidates in Fig. 3, and for each drug we also have conducted a rapid literature review^19^ for additional evidence:

**Pantoprazole** is a proton pump inhibitors (PPI) drug for treating gastroesophageal reflux disease (GERD), a damaged esophagus, and high levels of stomach acid caused by tumors. We observed that pantoprazole was associated with a 43% reduced risk of AD [hazard ratio (HR) 0.57, 95% confidence interval (CI) 0.56-0.59] in OneFlorida compared with a 8% reduced risk of AD (HR 0.92, 95% CI 0.89-0.94) in MarketScan. The association between using PPI drugs and risk of incident AD or non-AD dementias were contradictory^20,21^ in existing literature. Our study revealed one of the first large-scale RWD signals of pantoprazole for AD.

**Gabapentin** is an anti-epileptic drug for treating seizures and pain. We observed that gabapentin was associated with a 39% reduced risk of AD (HR 0.61, 95% CI 0.58-0.64) in OneFlorida and a 28% reduced risk of AD (HR 0.72, 95% CI 0.70-0.74) in MarketScan. Previous research suggested possible benefit of gabapentin for behavioural and psychological symptoms of dementia in AD patients based on summarizing case reviews^22^, and revealed crucial role of gabapentin in the Amyloid Beta Toxicity Cascade^23^. Our study showed one of the first large-scale RWD signals of gabapentin for AD.

**Acetaminophen** is used for treating mild to moderate pains and reducing fever. We observed that acetaminophen was associated with a 24% reduced risk of AD (HR 0.76, 95% CI 0.73-0.78) in OneFlorida and a 22% reduced risk of AD (HR 0.78, 95% CI 0.76-0.81) in MarketScan. Previous studies only indicated a weak association of acetaminophen with reduced risk of AD without any significance^24–26^ (e.g. Relative Risk 0.87, 95% CI 0.40-1.91 in a meta-analysis^24^).

**Atorvastatin** is used to treat high cholesterol and triglyceride levels, shows potentially beneficial but not significant effects on AD in^27,28^. We observed that atorvastatin was associated with a 21% reduced risk of AD (HR 0.79, 95% CI 0.77-0.81) in OneFlorida and a 12% reduced risk of AD (HR 0.88, 95% CI 0.85-0.90) in MarketScan.

**Albuterol** (also salbutamol) is a drug for asthma and chronic obstructive pulmonary disease (COPD). We observed that the albuterol was associated with a consistent 22% reduced risk of AD (HR 0.78, 95% CI 0.74-0.83) in OneFlorida and a 22% reduced risk of AD (HR 0.78, 95% CI 0.76-0.80) in MarketScan. Previous literature generated AD signals from in-vivo screening^29^ or rats models^30^. To the best of our knowledge, our work showed the first RWD signal of albuterol for AD.

**Fluticasone** is used to treat nasal symptoms, skin diseases, and also asthma. We observed that fluticasone was associated with a consistent 10% reduced risk of AD (HR 0.90, 95% CI 0.84-0.94) in OneFlorida and a 14% reduced risk of AD (HR 0.86, 95% CI 0.83-0.90) in MarketScan. Instead of high-throughput screening repurposing signals from RWD, Xu et al.^31^ validated fluticasone from MarketScan on a case by case basis and showed a consistent decreased risk for AD (HR 0.86, 95% CI 0.83–0.89) as ours, and Lehrer et al.^32^ also suggested a lower incidence of AD after taking fluticasone in another independent database, FDA MedWatch Adverse Events Database.

**Amoxicillin** is used to treat a wide variety of bacterial infections and stomach ulcers. We observed that amoxicillin was associated with a 12% reduced risk of AD (HR 0.88, 95% CI 0.82-0.95) in OneFlorida and a 7% reduced risk of AD (HR 0.93, 95% CI 0.90-0.96) in MarketScan. Jannis et al.^33^ revealed that the eradication of Helicobacter pylori (Hp) by a triple eradication regimen of omeprazole, clarithromycin and amoxicillin may positively influence AD manifestations in Hp-positive AD patients, and the action of different type of antibiotics in AD remains largely unknown^34^. To the best of our knowledge, our work revealed one of the first RWD signals of amoxicillin for AD.

**Omeprazole** is another PPI drug similar to pantoprazole. We observed that omeprazole was associated with a 12% reduced risk of AD (HR 0.88, 95% CI 0.85-0.91) in OneFlorida and a 8% reduced risk of AD (HR 0.92, 95% CI 0.88-0.94) in MarketScan. There is still no consensus on the role of PPIs and AD^20,21,35^. Our study showed one of the first large-scale RWD signals of omeprazole for AD.

## Discussion

We explored the problem of high-throughput clinical trial emulation on two large-scale RWD data warehouses, covering both EHRs and claims, in the context of identifying repurposable drug candidates for AD. There are several aspects we would like to highlight for our investigation.

- First, we emulated hundreds of trials for each drug based on two different ways of constructing control groups, which allowed for potentially more robust estimation of treatment effects. In our investigation, indeed, we observed a large variability (e.g., a large range of 95% confidence interval) of estimated treatment effects within emulated trials for certain drugs (e.g., Fig. 3e, albuterol FL-Rand, HR 0.78, 95% CI 0.70-0.86), and sometimes a large discrepancy between emulated trials when building control groups in different ways (Fig. 3f, fluticasone, FL-Rand, HR 0.75, 95% CI 0.70-0.81 versus FL-ATC, HR 1.01, 95% CI 0.97-1.05) These variabilities can become big challenges for existing observational studies that use a single control group^36^ or a single way of building multiple control groups (e.g. only random control groups)^7^.
- Second, we observed inconsistent results across the two data sets. For example, escitalopram showed a reduced risk in OneFlorida data (HR 0.61, 95% CI 0.55-0.67, Table. 1) but an increased risk in MarketScan database (HR 1.55, 95% CI 1.49-1.61, Table. 3). Potential explanations were rooted in intrinsic heterogeneity across the two datasets: OneFlorida is a regional database mainly covers patients’ EHRs in Florida area, while MarketScan is a nation-wide claims database across the US (Supplementary Tables 1). For example, the number of patients in escitalopram group in OneFlorida and MarketScan were 767 and 5,041 respectively. Such inconsistency highlights the necessity of leveraging at least two (different type of) data sets to derive robust and consistent evidence.
- Third, we conducted multiple sensitivity analyses to guarantee the robustness of our findings. We have investigated the impact of different ways of building control groups on balance performance (Supplementary Figs. 6). Our proposed model selection strategy greatly improved the performance of different PS models over conventional approaches. We also examined the influence of the balance diagnostics on the generated repurposing hypotheses. For example, if we adopted a more stringent balance criteria by requiring zero tolerance of unbalanced covariates (compared with 2% used in our primary analyses) in each emulated trial after re-weighting, we still recovered top four drugs among our reported eight drugs–pantoprazole (HR 0.60, 95% CI 0.57-0.63, OneFlorida; HR 0.92, 95% CI 0.89-0.94, MarketScan), gabapentin (HR 0.55, 95% CI 0.50-0.60, OneFlorida; HR 0.72, 95% CI 0.70-0.74, MarketScan), acetaminophen (HR 0.74, 95% CI 0.70-0.79, OneFlorida; HR 0.78, 95% CI 0.76-0.81, MarketScan), and atorvastatin (HR 0.78, 95% CI 0.75-0.81, OneFlorida; HR 0.88, 95% CI 0.85-0.90, MarketScan), which again significantly and consistently reduced risks for AD on both OneFlorida (Supplementary Table 4) and MarketScan (Supplementary Table 3) databases. We also studied different ways of constructing control groups–random controls and ATC-L2 controls (Method Section)–on the estimated treatment effects which were consistent for most of drugs (Fig. 3), except for fluticasone estimated from ATC-L2 controls on OneFlorida (Fig. 3f, FL-ATC, HR 1.01, 95% CI 0.97-1.05) and amoxicillin estimated from ATC-L2 controls on MarketScan (Fig. 3g, MS-ATC, HR 1.06, 95% CI 1.05-1.06).
- Last, compared with existing AD repurposing studies which typically focused on validating one or two hypotheses with a single type of RWD^31,37,38^, our study offered a high-throughput way of generating and validating AD repurposing hypotheses using both EHRs and claims^39^, which would further catalyze innovation in AD drug discovery at scale, or can be broadly applied to other diseases.

Lots of recent research efforts have been devoted to developing complex deep learning-based models for propensity score based modeling^7,40–43^In this paper, after emulating hundreds of thousands of trials from two large-scale RWD warehouses, we found that one LSTM-PS^7^, which is a representative deep learning based PS method, did not outperform LR-PS. Our study also highlighted the importance of model selection and we proposed our own strategy under which we demonstrated LR-PS outperformed gradient boosting tree-based PS models and deep multi-layer peceptron-based PS models as well in terms of balancing performance and the number of generated repurposing hypotheses. In addition, we also evaluated another model selection strategy widely used in literature^4,44–47^, which did not follow the out-of-sample validation strategy by partitioning data into complementary subsets but just estimated and evaluated PS model on the entire data set. We observed that with this strategy, even simple regularized LR-PS model suffered from over-fitting issue and could not be generalized well to unseen data in our empirical studies as demonstrated in Extended Data Fig. 5, emphasizing the need for a better model selection strategy for PS calculation. With all these investigations, we were able to show that our proposed model selection strategy, together with model training and evaluation pipelines, could serve as a better choice than existing model selection strategies for PS models in terms of goodness-of-balance and goodness-of-fit performance on emulated trials.

This study has several limitations. First, we identified MCI patients and AD onsets using ICD codes (Supplementary Tables 2) which were provided by physicians and validated in^48,49^, yet there might be a certain level of inaccuracy due to mis- and under-diagnosis or the lack of clinical details in EHRs or claims^39,50^. Information contained in clinical notes will be explored in the future through natural language processing to complement the structured codes. Second, although we balanced high-dimensional covariates collected during the baseline period, measurement error, residual confounding, and selection bias in the follow-up period were still possible. Therefore, adapting negative control^51^ for detecting residual confounding and selection bias to high-throughput trial emulation settings would be another promising direction.

## Conclusion

In this work, we proposed a high-throughput clinical trial emulation system for AD drug repurposing driven by RWD and propensity score-based causal inference with a tailored model selection strategy. On two large-scale RWD warehouses covering both EHRs and general claims, we demonstrated that our strategy identified eight drugs (pantoprazole, gabapentin, acetaminophen, atorvastatin, albuterol, fluticasone, amoxicillin, omeprazole) with different original indications could be potentially beneficial to AD patients. Our analyses highlighted model selection, rather than the PS model itself, is critical in balancing emulated trials at scale, which informs future RWD-based high-throughput trial emulation and can potentially accelerate the drug development process.

## Methods

### Data

We used two large-scale real-world longitudinal patient-level healthcare warehouses, including OneFlorida Clinical Research Consortium and IBM MarketScan Commercial Claims and Encounters (Data availability section). The OneFlorida database contains robust patient-level electronic health record (EHR) data for nearly 15 million (14,883,388) patients majorly from Florida and selected cities in Georgia and Alabama from January 2012 to April 2020, and the IBM MarketScan database (formerly known as Truven) contains administrative claim records from January 2009 to June 2020 for over 164 million (164,148,434) enrollees across the US, serving as a nationally representative database of the US population (See Supplementary Tables 1 for the population characteristics of two database). Both databases contain comprehensive longitudinal information on demographics, diagnoses, procedures, prescriptions, and outpatient dispensing for all enrollees. This study was approved by the Institutional Review Board of Weill Cornell Medicine with protocol number 21-07023759. The use of OneFlorida data for this study is approved under the University of Florida IRB number IRB202001888. Access to the MarketScan data analyzed in this manuscript is provided by the University of Kentucky.

### High-throughput trial emulation for Alzheimer’s disease (AD)

Instead of emulating one targeted randomized controlled trial on a case-by-case basis, here we tried to scale up trial emulation to a high-throughput setting, namely, to emulate hundreds of thousands of target trials to find potentially new indications of non-AD drugs for AD. We described the protocol of high-throughput trial emulations as follows and compared target trials and their emulations in the Extended Data Table 1. An illustration of the high-throughput cohort selection process was shown in Fig. 1a.

#### Eligibility criteria

We included patients with at least one mild cognitive impairment (MCI) diagnosis between January 2012 and April 2020 in the OneFlorida database (January 2009 to Jun 2020 in the MarketScan data). Other inclusion criteria were age at MCI diagnosis ≥ 50, no history of AD or AD-related dementia diagnoses before the baseline, the first MCI diagnosis date should be prior to the baseline, and ≥ 1 year of records before baseline. Of note, we defined the baseline as the first prescription date of the trial drug, and at baseline, all of the above criteria should have been met.

#### Treatment strategies

We compared two strategies for each drug trial: (0) no initiation of the trial drug before or after baseline (control group), and (1) initiation of the trial drug at baseline (treated group). We defined the treatment initiation date with the drug of interest as the first prescription date of the drug and we required at least two consecutive drug prescriptions over 30 days since the first prescription date in our database as a valid drug initiation.

#### Treatment assignment procedures

We classified patients into different drug groups according to their baseline eligibility criteria and their treatment strategies. We assumed the treated group and control group were exchangeable at baseline conditional on high-dimensional baseline covariates, including diagnoses, medications, demographics, and time from the MCI diagnosis date to drug initiation date. The diagnosis covariates consisted of selected comorbidities from Chronic Conditions Data Warehouse^52^ and established risk factors for AD selected by experts, resulting in 64 covariates; each defined by a set of selected ICD-9/10 codes. We grouped drug prescriptions coded as National Drug Code (NDC) or RXNORM codes into their major active ingredients coded in RXNORM defined in Unified Medical Language System^53^ for the OneFlorida case, and into the Medi-Span Generic Product Identifier (GPI)^54^ by their first 8 digits for the MarketScan data. We used the first 200 most commonly prescribed drug ingredients for the co-prescribed medication covariates for each drug trial and thus the medication covariates varied in different drug trials. We used 2 covariates age and sex for demographics and 1 covariate for the time from the MCI diagnosis date to the drug initiation date. In total, there were 267 covariates to adjust for. In addition to the 267 baseline covariates, we also considered the temporal sequences of each of diagnoses and medications for the deep long short-term memory network with attention mechanisms-based PS calculation^7^.

#### Follow-up

We followed each patient from his/her baseline until the day of the first AD diagnosis, loss to follow-up (censoring), 2 years after baseline, or the end date of our databases, whichever came first.

#### Outcomes

The outcome of interest is the diagnosis of AD recorded in the database within his/her follow-up period, which was denoted as a positive event. If there was no AD diagnosis recorded in a patient’s follow-up period, and the last prescription date or the last diagnosis date recorded in the database came after the end of the follow-up, then we marked it as a negative event. A censoring event is a case where there was no AD diagnosis recorded in a patient’s follow-up period and the last prescription date and the last diagnosis date recorded in the database came before the end of the follow-up. The time to positive event is defined as the days between the baseline date and the first diagnosis of AD. The time to negative event is the time of follow-up. The time to censoring is defined as the days between the baseline date and the last prescription date or the last diagnosis date, whichever comes last. Clinical phenotypes were identified by the selected diagnosis codes by experts (Supplementary Tables 2).

#### Causal contrasts of interest

The observational analogy of intention-to-treat effect of being assigned to trial drug initiation versus no initiation at baseline.

#### High-throughput emulation

We emulated trials for all drugs appeared in our databases with at least 500 eligible patients in their treated groups. For each emulated trial, its treated group consists of eligible patients who initiated the trial drug, and its control group consists of eligible patients who had no initiation of the trial drug. We constructed the no-initiation patients group in two ways: a) randomly selecting eligible patients from other drug initiation group^49^, or selecting patients from similar drug groups that are under the same second-level Anatomical Therapeutic Chemical classification category^55^ (ATC-L2) as the target trial drug^6^. We further excluded any of those patients who were also in the trial drug group or prescribed the trial drug before baseline. To investigate statistically significance of results with varying control groups, we emulated 100 trials for each targeted drug and among which 50 emulated trials adopted random controls and the other 50 emulated trials adopted ATC-L2 controls as described above. Different combinations of control groups were studies as sensitivity analysis.

### Causal effect estimation and the screening of repurposing drugs

We used propensity score (PS) methods^56^ for confounding control and treatment effect estimation for high-throughput emulated trials, and proposed two criteria to screen and prioritize non-AD drugs for repurposing (Summarized in Fig. 1b).

#### Propensity score and IPTW

For each emulated trial, we used propensity score (PS) framework^56^ to learn empirical treatment assignment given baseline covariates, and used the inverse probability of treatment weighting (IPTW)^57^ to balance treated and control groups. We used triplet (*X, Z,Y, T*) to represent data of both treated and control groups where *X, Z, Y, T* represent the baseline covariates, treatment assignment, outcome indicator, and time to events, respectively. The PS is defined as *P*(*Z* = 1|*X*)^56^ where *Z* is treatment assignment (*Z* = 1 and *Z* = 0 for treated and control respectively) and *X* denotes patients’ observed baseline covariates. The inverse probability of treatment weight (IPTW) is defined as 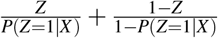^57, 58^, which tries to make original trial into a more balanced pseudo trial by re-weighting each data sample. We used an updated version named stabilized IPTW, defined as

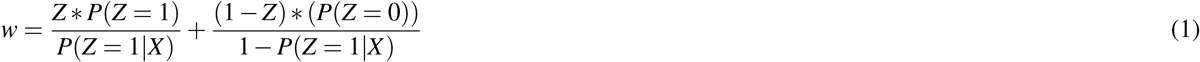

to deal with extreme re-weighting weights and thus potentially inflated sample size^7,59,60^.

A machine learning (ML) or deep learning (DL)-based propensity score (ML/DL-PS) model is a binary classification model *f*_*θ*_ ∈ *ℱ*_Θ_ : *X* → *Z*, to approximate *P*(*Z* = 1 |*X*) by *f*_*θ*_ (*X*) with learnable parameters *θ*. Here, we use *ℱ*_Θ_ to denote a set of ML/DL models (e.g. a set of models with varying hyper-parameters) and *f*_*θ*_ to denote one specific model instance in this set. We considered four classes of ML/DL *ℱ*_Θ_: (a) regularized logistic regression-based PS models (LR-PS), encompassing its special case logistic regression (without any regularization term), which are most widely used model for PS calculation; (b) the state-of-the-art deep learning based-PS model, long short-term memory network with attention mechanisms-based PS models (LSTM-PS)^7^; c) multi-layer perception network-based PS models (MLP-PS)^12,13^; and d) the state-of-the-art gradient boosted tree-based PS models (GBT-PS)^9–11^.

#### Performance evaluation criteria

We evaluated the performance of estimated PS models in terms of two aspects: a) the goodness-of-balance, and b) the goodness-of-fit.

The goodness-of-balance is measured by the standardized mean difference (SMD)^17,44,61^ on the whole dataset, defined as follows:

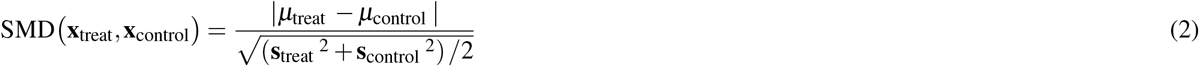

where **x**_treat_, **x**_control_ ∈ **R**^*D*^ represent the vector representations of *D* covariates of treated group and control group respectively, *µ*_treat_, *µ*_control_ ∈ **R**^*D*^ are their sample means over the treated group and control group respectively. Similarly, **s**_treat_^2^, **s**_control_^2^ ∈ **R**^*D*^ are their sample variances. Suppose that we have learned sample weight *w*_*i*_ for each patient *i* by IPTW, the weighted sample mean and variance are:

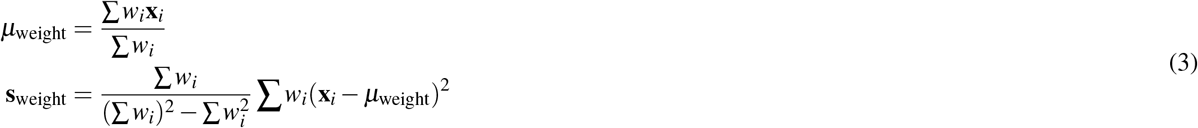

The weighted versions of sample mean and variance hold for both treated and control groups and thus we ignored their corner marks for brevity. The SMD_weight_ can be calculated by applying above weighted mean and variance to Eq.2. All operations in Eq.2 and 3 are conducted in an element-wise way for each covariate. For each dimension *d* of either original SMD or weighted SMD, it is considered balanced if its *d*^*th*^ SMD value SMD(*d*) ≤ 0.1^17^, and the treated and control groups are balanced if the total number of unbalanced features ≤ 2%∗*D*^7^. More stringent balance criteria (e.g., requiring non-unbalanced features) were also considered as sensitivity analysis. Taking IPTW re-weighted case as an example, we can calculate the number of balanced feature after IPTW by:

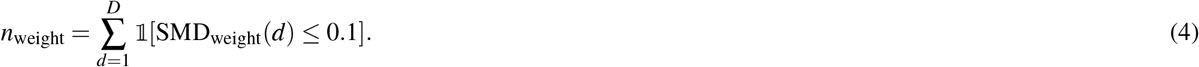

The smaller the *n*_weight_ is, the better the balance performance of IPTW is, and the less biased estimated causal effect is. As shown in^61^, SMD is one of the top predictors of the bias of estimated causal effect. To quantify balance performance of high-throughput emulation of one drug trials, we further defined the probability of successfully balancing one specific drug *M* trial by a set of PS models *ℱ*_Θ_ as 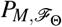, which can be estimated by the fraction of successfully balanced trials over all emulations as follows:

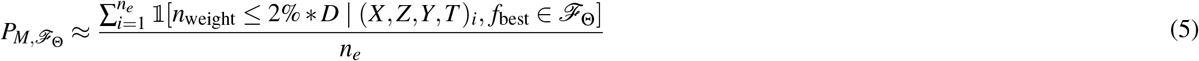

where *n*_*e*_ is the total number of emulated trials (*X, Z,Y, T*)_*i*_, *i* = 1, 2, …, *n*_*e*_ for drug *M, f*_best_ is the best PS model among *ℱ*_Θ_ learned from the *i*^*th*^ emulated trial, and the IPTW and *n*_weight_ are calculated by applying *f*_best_ to the *i*^*th*^ emulated trial. We will discuss how to learn and select *f*_best_ ∈ *ℱ*_Θ_ in the next section. In general, the larger the balancing success rate *P*_*M*_(*n*_weight_ ≤ 2% ∗*D*|*ℱ*_Θ_) is, the better the *ℱ*_Θ_ model balances the drug *M* trial.

The goodness-of-fit is the generalized prediction performance of the PS model on the unseen data. We used the area under the receiver operating characteristic (AUC) measured on the (unseen) testing dataset to quantify it^62,63^. The larger AUC on the testing set is, the better the generalization performance of the classification model is.

#### Model training, selection and evaluation

Much existing literature used statistical models (e.g. logistic regression) for PS calculation in estimating treatment effect from observational data, and the PS model was both estimated from and applied to the whole dataset^4,44–47^. By contrast, machine learning or deep learning (ML/DL) models, which are good at capturing complex data and usually have a large number of hyper-parameters, are faced with over-fitting and generalization trade-off^64^. Thus, to get more generalized ML/DL models, the conventional approach is to split the whole dataset into complementary training, validation, and testing sets; then train the model on the training set, select the best learned model according to the AUC performance for example (a goodness-of-fit measure; the larger the better) on the validation set, and finally evaluate the selected model on the testing set. This model validation strategy for ML/DL models is also known as the (one-round) cross-validation^14^. Following this conventional data splitting strategy, existing ML or DL-based PS models^9–13,49^ selected the best model by the AUC (a goodness-of-fit measure; the larger the better) or the maximum SMD value (a goodness-of-balance measure; the smaller the better) measured on the validation set. However, following the above model selection strategies for PS model selection for balancing emulated trials, poor balance performance was observed in our high-throughput study in both two RWD warehouses.

Here, we introduce our model training and selection algorithm tailored for ML/DL-based PS model in Algorithm 1, trying to get the best goodness-of-balance performance as well as the best possible goodness-of-fit performance. We used binary cross-entropy loss *ℒ* as the objective function for learning empirical binary propensity scores. We also describe the evaluation (testing) algorithm for ML/DL-based PS models in Algorithm 2, to evaluate and benchmark different learned and selected models. Of note, the goodness-of-balance is the single most important criterion for evaluating trial emulations. If the balancing results are tied, the goodness-of-fit is evaluated.

#### Statistical analysis

We reported adjusted 2-year survival difference by adjusted Kaplan–Meier estimator^65,66^ and adjusted hazard ratio (HRs) modeled by the adjusted Cox proportional hazard model^66,67^ for each of the emulated trials. The above outcome estimators were adjusted by inverse probability of treatment weighting (IPTW) based on the best PS calculation selected by our model selection strategy. For each drug we reported their sample means of different outcome estimators with 95% confidence intervals^68^ over all the balanced trials. We used two ways of building different control groups (e.g., random controls or ATC-L2 controls) and different balance criteria (e.g., different thresholds for SMD) to evaluate the robustness of our estimated effects in various sensitivity analyses.

##### Algorithm 1 ML/DL-PS model training and selection algorithm

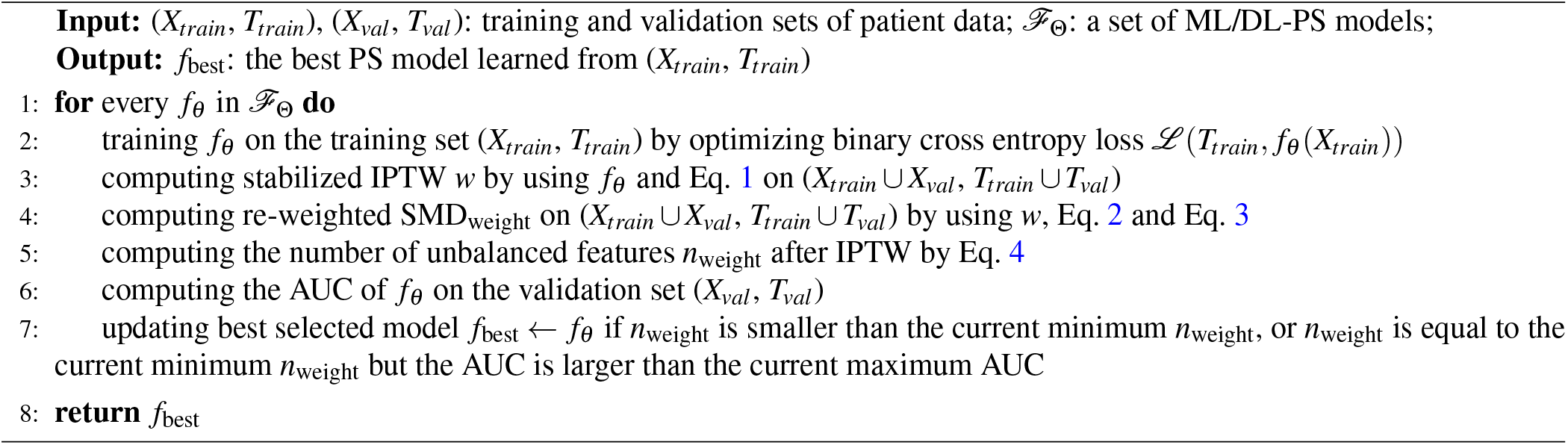

##### Algorithm 2 ML/DL-PS model evaluation (testing) algorithm

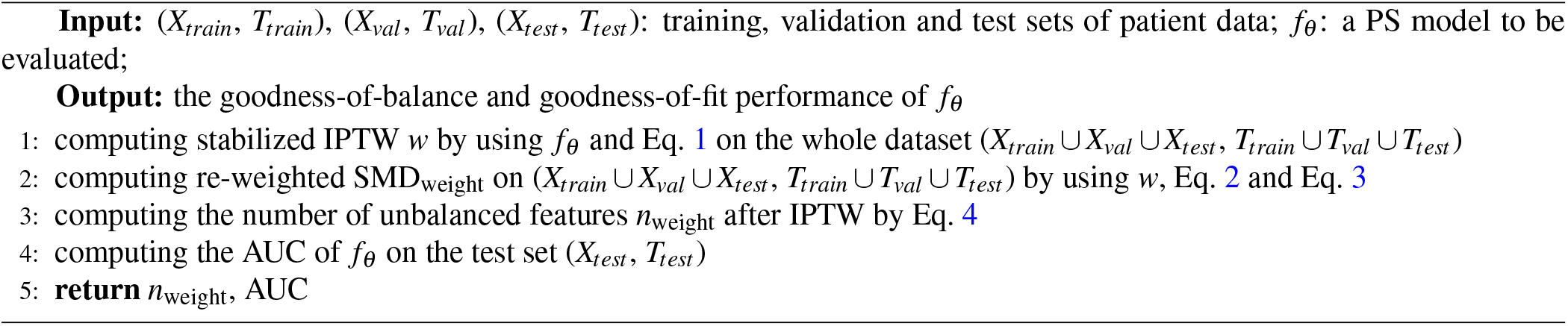

#### Screening and prioritization

To generate reliable and robust repurposing hypotheses for AD, we required that the estimated effects of repurposing drug candidates should be significantly and consistently beneficial. As for the significant (beneficial) effects, we require that the fraction of successfully balanced trials of a drug candidates after IPTW ≥ 10%, and their adjusted 2-yr survival difference of these balanced trials should be significant. We used bootstrapping hypothesis testing^68^ to test if the sample mean of the adjusted 2-yr survival difference from all the balanced trials is *>* 0 (*<* 1 for HRs), and we considered p-value *<* 0.05 as significant. As for the consistency of effects, we required that the estimated effects should be all significantly beneficial over different databases. We then ranked the drug candidates according to their estimated effects. More stringent screening criteria were considered in sensitivity analysis.

#### Comparison with existing works

We replicated the analytic approach by Liu et al.^7^ and we found that their methods led to biased SMD estimation and worse balance performance as shown in Table 1 due to their deep LSTM-based PS methods. Besides, there are other major concerns. First, they selected patients at baseline according to patients’ treatment strategy over follow-up and such post-baseline information should not be used at baseline^46^. Second, they estimated treatment effect by the average treatment effect (ATE) ATE = 𝔼[*Y*_1_ − *Y*_0_] (*Y*_1_ and *Y*_0_ are the potential outcomes for each patient under the treatment or the control respectively), which can introduce selection bias due to loss to follow-up (censoring)^69^. Third, they generated hypotheses only on one database and used only random controls, ignoring the potential variability we found over different databases and over emulations with different control groups.

#### Experimental settings

We implemented our high-throughput clinical trial emulation system for drug repurposing by Python 3.9 and Pytorch 1.8 (https://pytorch.org/ and trained deep learning models by Adam optimizer^70^ on a Linux server with two GeForce RTX 2080 Ti GPUs and 16 CPU cores. We used python package lifelines-0.26^66^ for survival analysis, scikit-learn-0.23^18^ for machine learning models including regularized logistic regression, and lightgbm-3.2^11^ for gradient boosted machine. We followed Liu et al.^7^ for their LSTM-PS implementations. We randomly partitioned each emulated trial into complementary training, validation and testing data sets with a ratio of 70:10:20. Please refer to our python package for more details.

## Data Availability

OneFlorida data can be requested through https://onefloridaconsortium.org/front-door/. Since OneFlorida data is a HIPAA-limited data set, a data use agreement needs to be established with the OneFlorida network.The MarketScan dataset is available from IBM at https://www.ibm.com/products/marketscan-research-databases.

## Code availability

For reproducibility, we open-sourced our python code package at https://github.com/calvin-zcx/RWD4Drug.

## Data Availability

The OneFlorida data can be requested through https://onefloridaconsortium.org/front-door/. Since the OneFlorida data is a HIPAA-limited data set, a data use agreement needs to be established with the OneFlorida network. The MarketScan dataset is available from IBM at https://www.ibm.com/products/marketscan-research-databases.

## Acknowledgements

The work of C.Z., H.Z. and F.W. were supported by NIH awards R01MH124740 and RF1AG072449, as well as NSF award 1750326. B.S.G. is supported by NIH grant RF1AG059319. J.B. is supported by NIH awards R21AG068717, R56AG069880, R01CA246418-02S1, and R21CA245858-01A1S1.

## Author contributions statement

CZ and FW proposed the initial idea. CZ designed and implemented the framework, analyzed the data, and wrote the initial draft of the paper. Hansi Zhang, JX and JB processed and analyzed the OneFlorida data. SF and JC processed the data and ran the experiments on the MarketScan database. Hao Zhang, SH and BSG helped with the data analysis and result interpretation. KC and YC designed the statistical analysis. All the authors contributed to the discussions of results and final writing of the paper. JC had access to the raw data of the MarketScan database. JB had access to the raw data of the OneFlorida database.

---

**Extended Data Table 1.** **A summary of the protocol of target trials and high-throughput emulations to estimate the effect of drugs on AD risk using real-world healthcare data OneFlorida (2012-2020) and MarketScan (2009-2020).**

| Protocol component | Target trial specification | Target trial emulation |
| --- | --- | --- |
| Eligibility criteria* | Patients with MCI between January 2012 and April 2020 in OneFlorida database (January 2009 and June 2020 in MarketScan database)<br>Age $\geq 50$ at MCI diagnosis, and no upper age limit.<br>No history of AD or dementia before baseline<br>No trial drug prescription before baseline<br>Baseline is defined as date when all eligibility criteria are met.<br>*The criteria were adapted from trail NCT00151502 for the settings of high-throughput emulation on RWD. | Same as for the target trial<br>We defined MCI diagnosis according to the selected ICD-9/10 codes in patients' baseline period to identify probable and potential mild AD cohorts<br>We required $\geq 1$ year from one individual's first record in database to his/her baseline<br>We required the first MCI diagnosis before baseline<br>Baseline is defined as the first date of the trial drug prescription and at that time point all eligibility criteria are met |
| Treatment strategies | Strategy 0: No initiation of any trial drug before or after baseline.<br>Strategy 1: Initiation of trial drug at baseline. | Same as for the target trial<br>We defined the medication initiation date to be the first date of a prescription of the trial drug and we require $\geq 2$ prescriptions within $\geq 30$ days from the initiation date as a valid initiation. |
| Treatment assignment | Patients are randomly assigned to either treatment strategy at baseline and are aware of the strategy they are assigned to. | We classified patients into different groups according to their baseline eligibility criteria and treatment strategy.<br>We assumed that the treated group and control group were exchangeable by adjusting for high-dimensional confounders collected before the baseline, including: demographics, diagnoses, medications, time lag between MCI initiation and index date, etc. |
| Outcomes | Diagnosis of AD | Same as for the target trial, we defined the AD diagnosis according to selected ICD-9/10 codes in the follow-up period. |
| Follow-up | We followed each patient from his/her baseline date until the date of his/her first AD diagnosis, loss to follow-up, or 2 years (730 days) after the baseline, whichever happens first. | Same as for the target trial |
| Causal contrast | Intention-to-treat effect | Observational analog of intention-to-treat effect |
| High-throughput trials | For each drug among a large number of drug candidates, we conducted a target trial following the above protocol to estimate its effect. | We emulated trials for all the drugs in our database with $\geq 500$ patients in the trial drug group, and for each drug we emulated 100 trials by constructing different control groups as follows.<br>For each emulated trial, the treated group consists of patients who were eligible and adopted the trial drug strategy according above protocol, and its control groups consist of eligible patients either from randomly chosen drug groups other than the trial drug group, or from similar drug groups within the same second level ATC category as the trial drug, and we further excluded any of them who were also in the trial drug group or who prescribed the trial drug before baseline. |
| Statistical analysis | Intention-to-treatment analysis as the time-to-first event<br>Applying IPTW to adjust for baseline confounders<br>Non-parametric bootstrapping for 95% CIs | Same intention-to-treat analyses. Applying different ML/DL-based PS models to adjust for high-dimensional baseline covariates by IPTW. The best PS model was selected by our proposed ML/DL-PS model selection strategy.<br>Adjusted 2-yr survival difference by adjusted KM method, and adjusted HRs by adjusted CoxPH, and we report sample means with 95% bootstrapped CIs for balanced trials from high-throughput emulations<br>Sensitivity: estimated effects by building different control groups (random controls or ATC-L2 controls), and by different balance criteria. |

MCI, mild cognitive impairment; AD, Alzheimer's disease; KM, Kaplan-Meier; HR, hazard ratio; CoxPH, Cox proportional hazards; CIs, confidence intervals; ML/DL, machine learning or deep learning; IPTW, inverse-probability treatment weights; PS, propensity score

**Extended Data Fig. 1.**
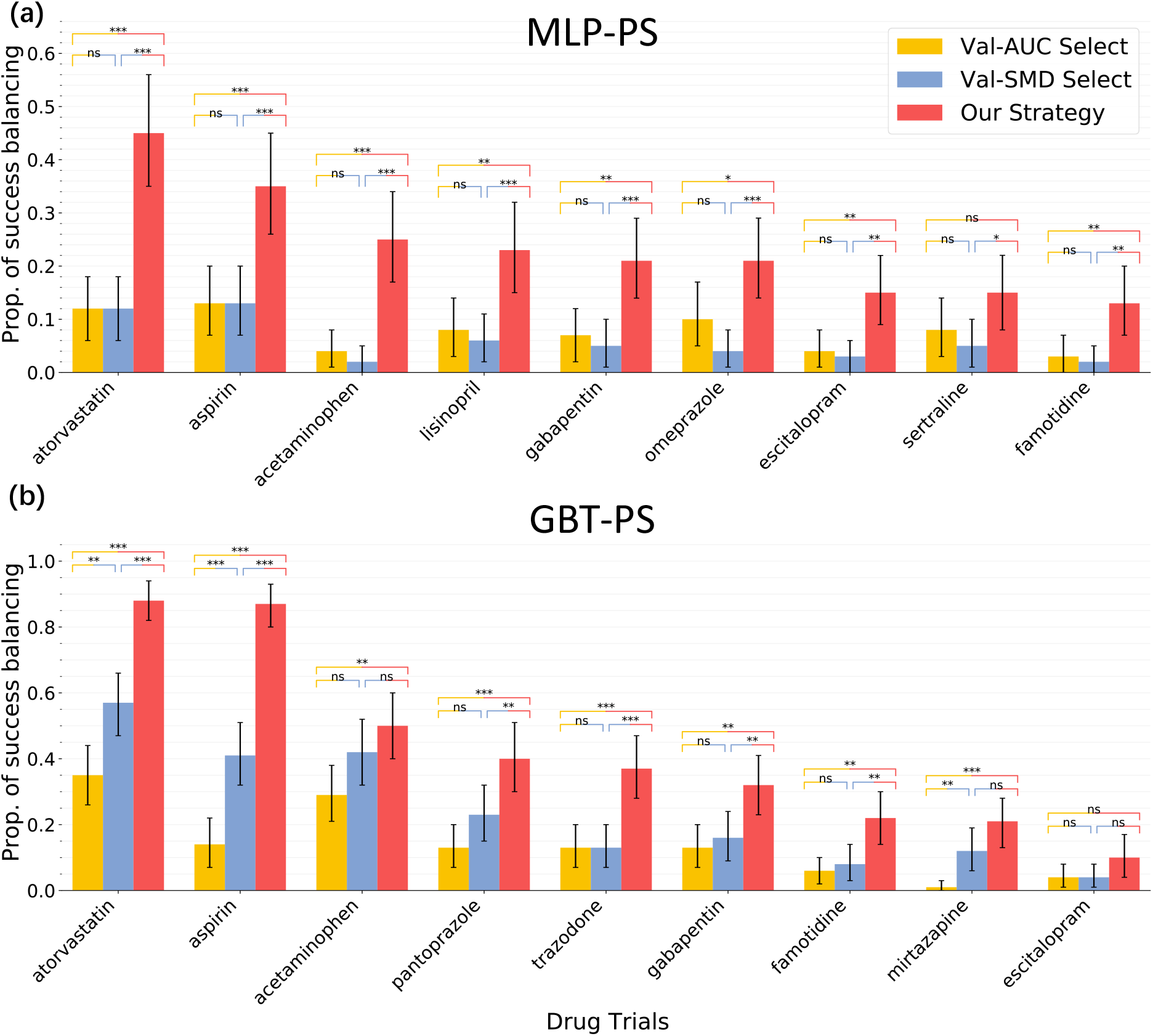
Proportion of successfully balanced drug trials by (a) MLP-PS and (b) GBT-PS models selected under different model selection strategies, OneFlorida database, 2012-2020. Propensity score models selected by our model selection strategy balanced significantly more trials than other model selection methods for all target drugs. We reported drug trials with at least 10% balanced trials based on 100 emulated trials for each drug. The error bars mean 95% confidence intervals by 1000-times bootstrapping. The (two-sided) independent two-samples T-test for testing the means of each two bars, and *, *p <* 0.05; **, *p <* 0.01; ***, *p <* 0.001; not significant (ns), *p ≥* 0.05; MLP-PS, multi-layer perceptron-based propensity score models; GBT-PS, gradient boosted tree-based propensity score models.

**Extended Data Fig. 2.**
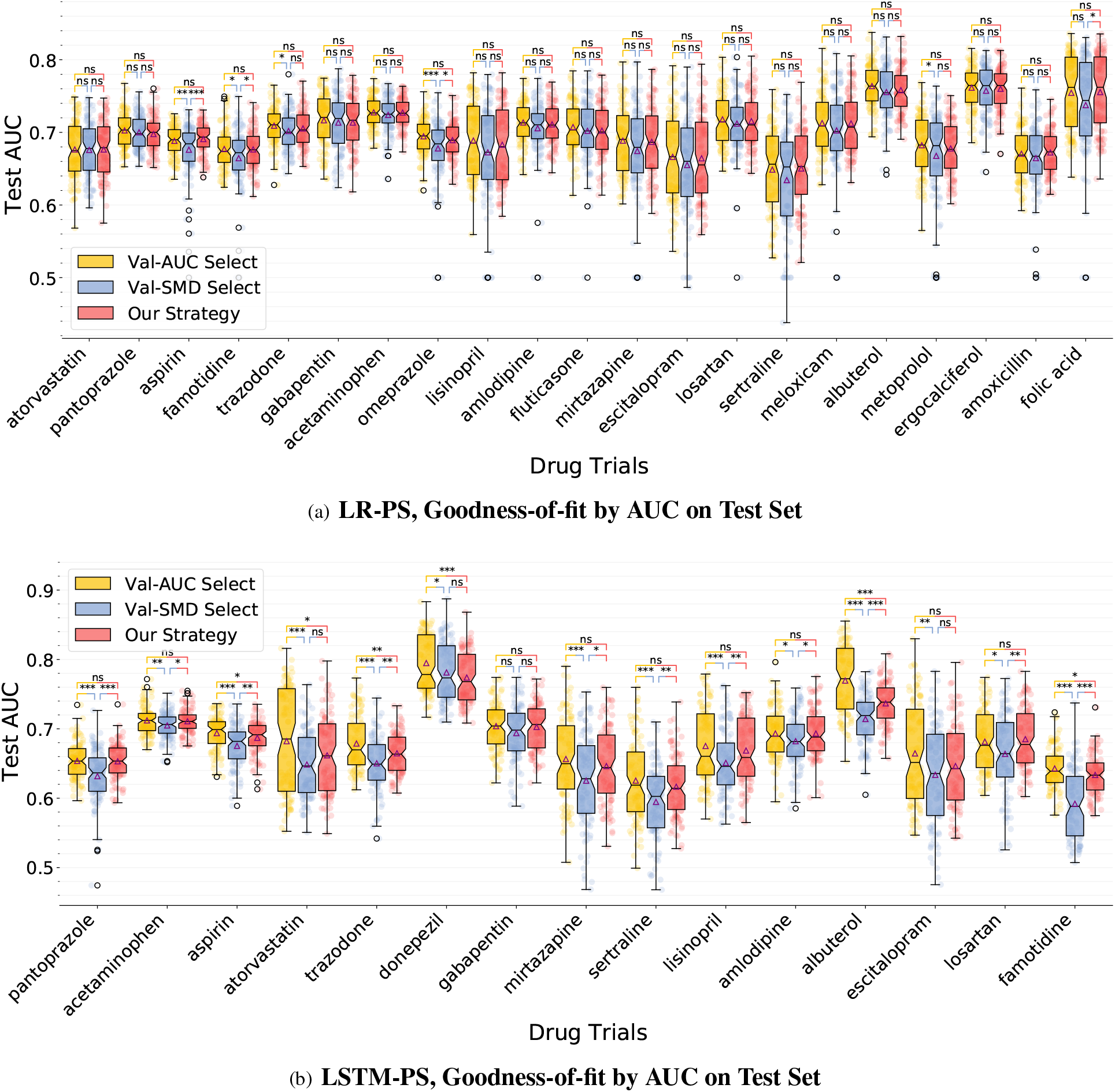
Distribution of AUC performance on (unseen) test data by (a) LR-PS and (b) LSTM-PS models selected under different model selection strategies, OneFlorida database, 2012-2020. We reported drugs with at least 10% balanced trials based on 100 emulated trials for each drug. Box plots with 25th (Q1, lower quartile), median (central vertical line), 75th (Q3, upper quartile), and whiskers extending to 1.5 interquartile range (IQR=Q3-Q1). Triangle marks represent sample means. The (two-sided) independent two-samples T-test for testing the means of each two bars, and *, *p <* 0.05; **, *p <* 0.01; ***, *p <* 0.001; not significant (ns), *p ≥* 0.05; AUC, the area under the receiver operating characteristic curve; LR-PS, regularized logistic regression-based propensity score models; LSTM-PS, long short-term memory network with attention mechanisms-based propensity score models.

**Extended Data Fig. 3.**
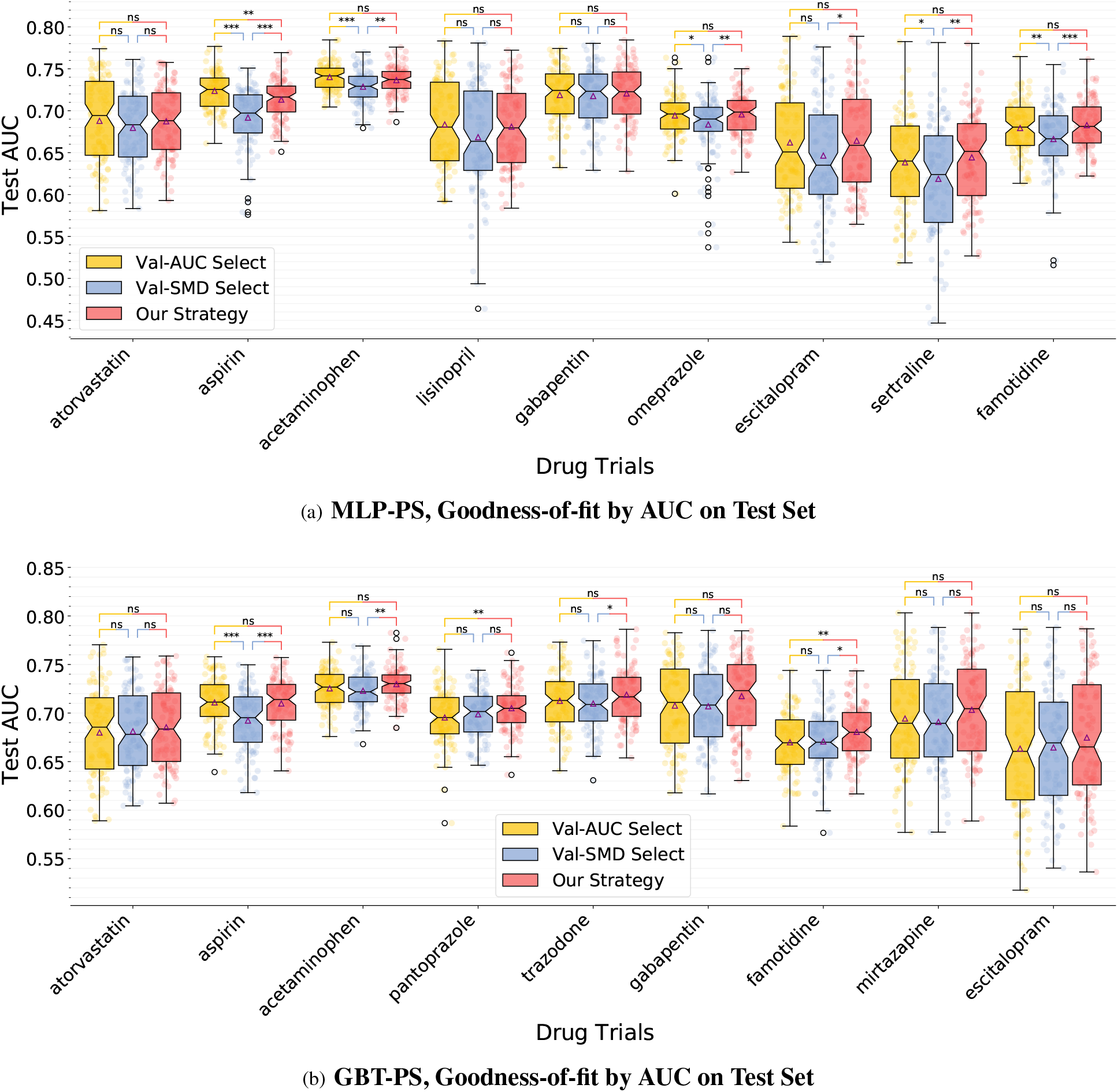
Distribution of AUC performance on (unseen) test data by (a) MLP-PS and (b) GBT-PS models selected under different model selection strategies, OneFlorida database, 2012-2020. We reported drugs with at least 10% balanced trials based on 100 emulated trials for each drug. Box plots with 25th (Q1, lower quartile), median (central vertical line), 75th (Q3, upper quartile), and whiskers extending to *±*1.5*×* interquartile range (IQR=Q3-Q1). Triangle marks represent sample means. The (two-sided) independent two-samples T-test for testing the means of each two bars, and *, *p <* 0.05; **, *p <* 0.01; ***, *p <* 0.001; not significant (ns), *p ≥* 0.05; AUC, the area under the receiver operating characteristic curve; MLP-PS, multi-layer perceptron-based propensity score models; GBT-PS, gradient boosted tree-based propensity score models.

**Extended Data Fig. 4.**
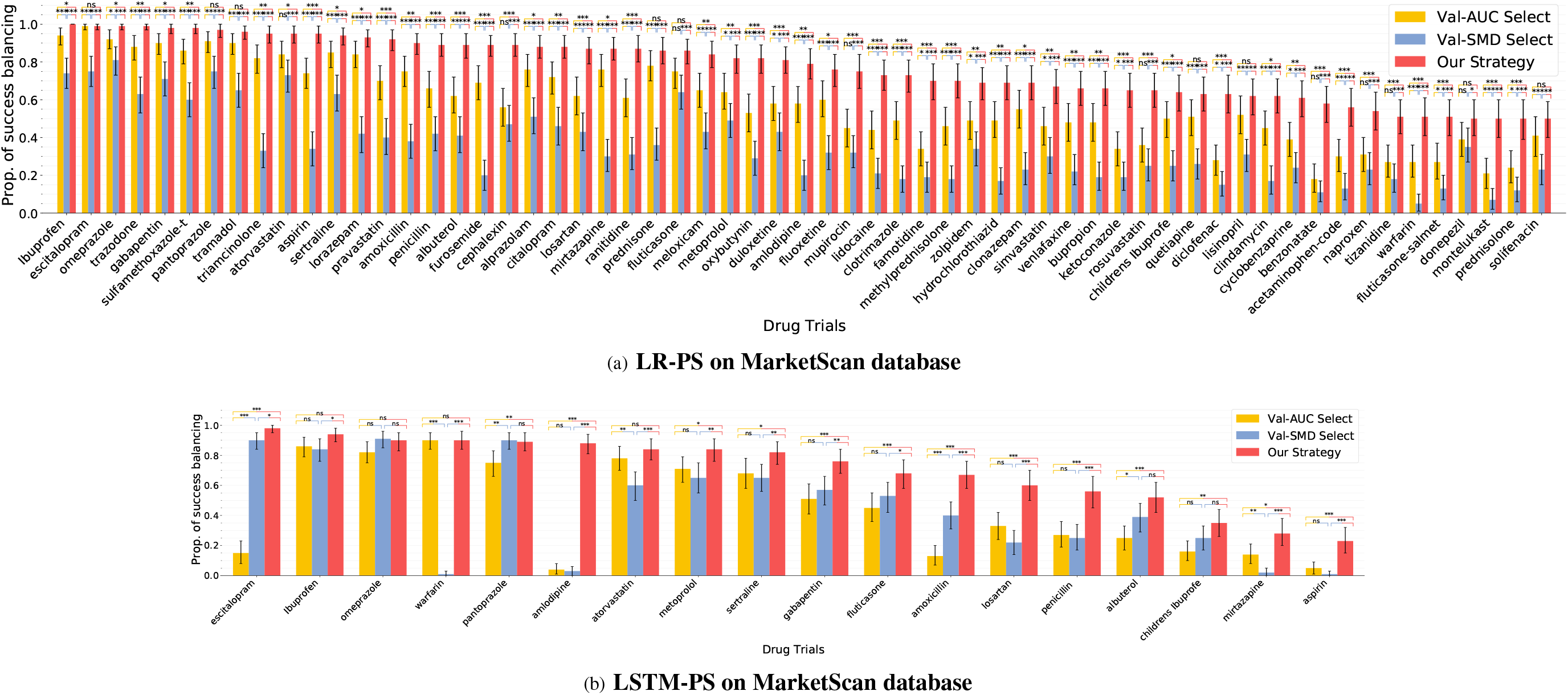
Proportion of successfully balanced drug trials by (a) LR-PS and (b) LSTM-PS models selected under different model selection strategies, MarketScan database, 2009-2020. PS models selected by our model selection strategy balanced significantly more trials than other model selection methods for all target drugs. We applied LR-PS to all drug trials with ≥500 treated patients in MarketScan and we reported drug trials among which 50% trials were balanced after re-weighting based on 100 emulations trials. We applied LSTM-PS to drug candidates selected by LR-PS and reported drugs with 10% balanced trials because LSTM-PS is not scalable to all existed drugs in MarketScan as in LR-PS case. We required one re-weighted trial to be balanced if all high-dimensional covariates were balanced after IPTW. The error bars mean 95% confidence intervals by 1000-times bootstrapping. The (two-sided) independent two-samples T-test for testing the means of each two bars, and *, *p <* 0.05; **, *p <* 0.01; ***, *p <* 0.001; not significant (ns), *p ≥* 0.05; LR-PS, regularized logistic regression-based propensity score models; LSTM-PS, long short-term memory network with attention mechanisms-based propensity score models^7^; IPTW, inverse-probability treatment weights; PS, propensity score.

**Extended Data Fig. 5.**
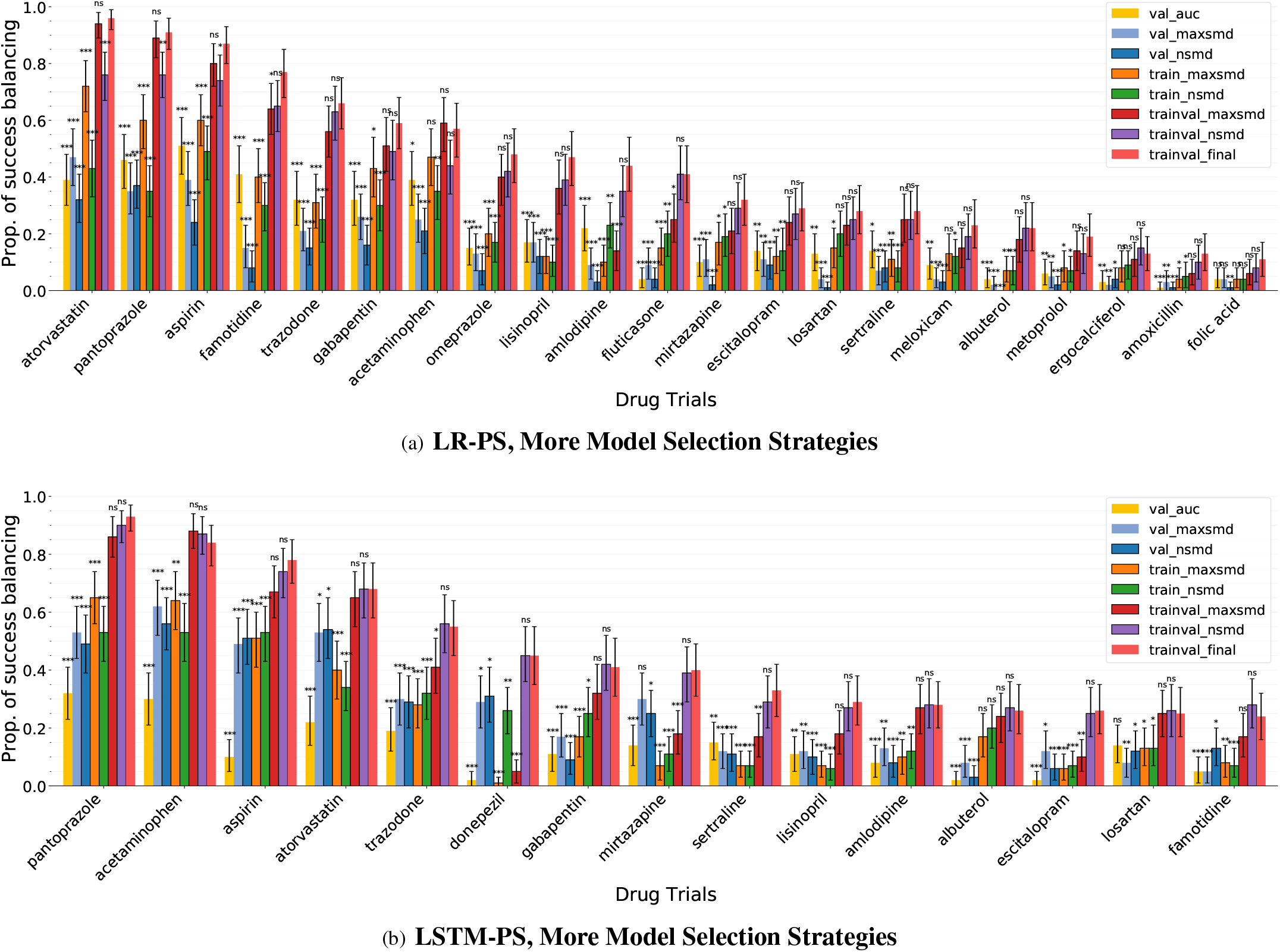
Proportion of successfully balanced drug trials by (a) LR-PS and (b) LSTM-PS models selected under more model selection strategies, OneFlorida database, 2012-2020. Propensity score models selected by our model selection strategy balanced significantly more trials than other model selection strategies. Different color bars denoted balancing performance on the whole data by the best PS model selected under different model selection strategies, including: (1) val_auc, model selection by AUC score on the validation set; (2) val_maxsmd, by maximum SMD after IPTW on the validation set; (3) val_nsmd, by the number of unbalanced feature after IPTW on the validation set; (4) train_maxsmd, by the maximum SMD after IPTW on the training set; (5) train_nsmd, by the number of unbalanced feature after IPTW on the training set; (6) trainval_maxsmd, by the maximum SMD after IPTW on the training and validation combined set; (7) trainval_nsmd, by the number of unbalanced feature after IPTW on the training and validation combined set; (8) trainval_final, our model selection strategy based on both the number of unbalanced feature after IPTW on the training and validation combined set and AUC score on the validation set. We reported drug trials with at least 10% balanced trials based on 100 emulated trials for each drug. The error bars mean 95% confidence intervals by 1000-times bootstrapping. The (two-sided) independent two-samples T-test for testing the difference between each method versus our final strategy, and *, *p <* 0.05; **, *p <* 0.01; ***, *p <* 0.001; not significant (ns), *p ≥* 0.05; LR-PS, regularized logistic regression-based propensity score models; LSTM-PS, long short-term memory network with attention mechanisms-based propensity score models^7^; IPTW, inverse-probability treatment weights; PS, propensity score.

**Extended Data Fig. 6.**
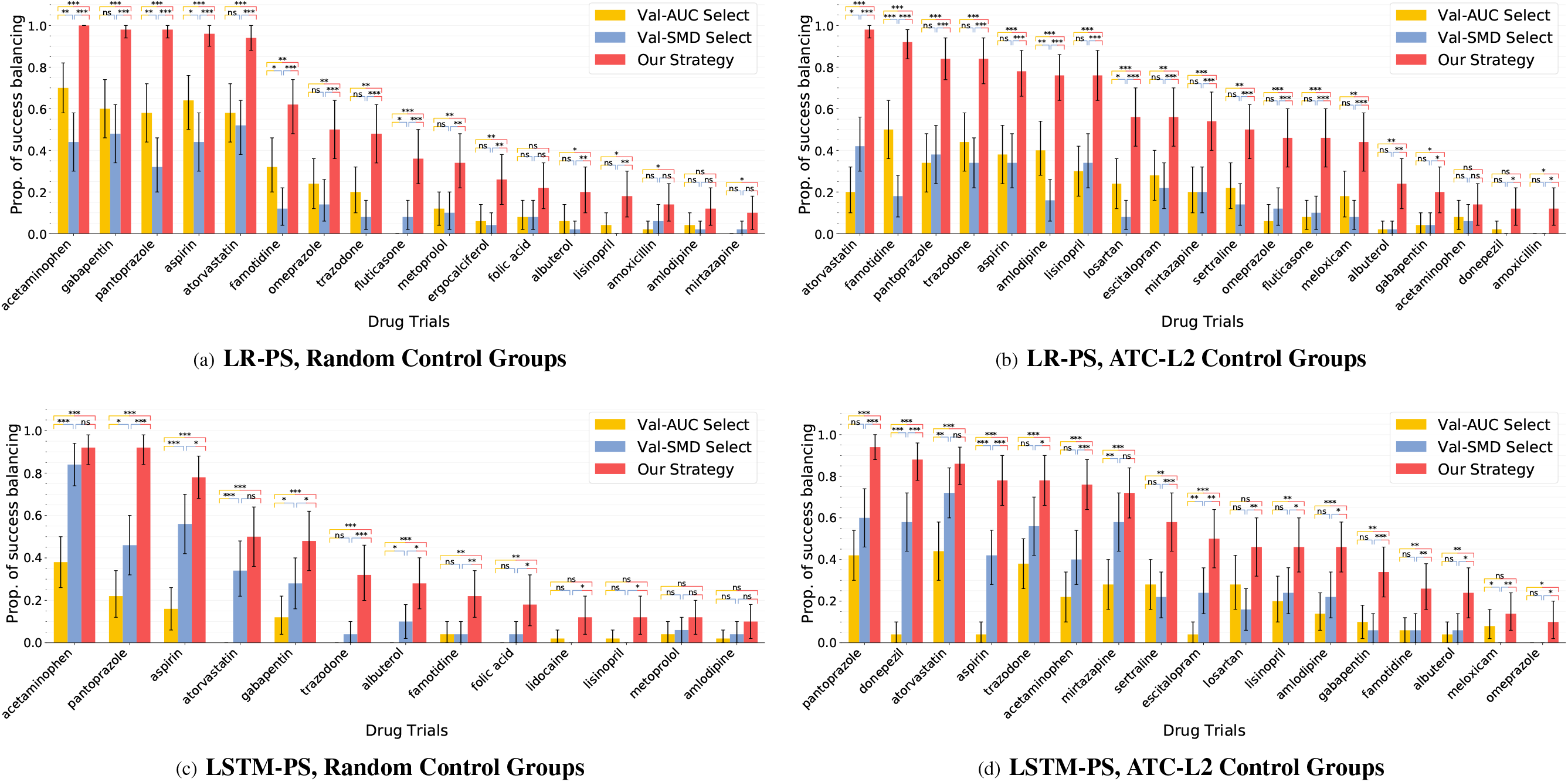
Proportion of successfully balanced drug trials by (a) LR-PS and (b) LSTM-PS models selected under different model selection strategies and different ways of emulating control groups, OneFlorida database, 2012-2020. Propensity score models models selected by our model selection strategy balanced significantly more trials than other model selection methods for all target drugs. We reported drug trials with at least 10% balanced trials based on 100 emulated trials for each drug. The error bars mean 95% confidence intervals by 1000-times bootstrapping. The (two-sided) independent two-samples T-test for testing the means of each two bars, and *, *p <* 0.05; **, *p <* 0.01; ***, *p <* 0.001; not significant (ns), *p ≥*0.05; LR-PS, regularized logistic regression-based propensity score models; LSTM-PS, long short-term memory network with attention mechanisms-based propensity score models^7^.

## Supplemental Materials for

**Supplementary Table 1.** Characteristics of base population in the two real-world healthcare databases for high-throughput trial emulation.

| **OneFlorida, January 2012 to April 2020, 14,883,388 patients** |  |  |  |  |
| --- | --- | --- | --- | --- |
|  | **MCI** | **AD** | **MCI \ AD** | **P-value<sup>a</sup>** |
| No. of patients | 73,927 (100%) | 10,530 (14.24%) | 63,397 (85.76%) | – |
| MCI age, median (IQR) <sup>c</sup> | 66 (50, 77) | 78 (71, 85) | 64 (46, 75) | 0.000 |
| Sex-female | 40,654 (54.99%) | 6,681 (63.45%) | 33,973 (53.59%) | 0.000<sup>b</sup> |
| Sex-male | 33,273 (45.01%) | 3,849 (36.55%) | 29,424 (46.41%) | – |
| Antidiabetic medication | 19,307 (26.12%) | 2,563 (24.34%) | 16,744 (26.41%) | 0.000 |
| Antihypertensives medication | 10,909 (14.76%) | 1,208 (11.47%) | 9,701 (15.30%) | 0.000 |
| Alcohol Use Disorders | 7,728 (10.45%) | 709 (6.73%) | 7,019 (11.07%) | 0.000 |
| Anxiety Disorders | 38,602 (52.22%) | 6,009 (57.07%) | 32,593 (51.41%) | 0.000 |
| Depression | 41,763 (56.49%) | 6,979 (66.28%) | 34,784 (54.87%) | 0.000 |
| Diabetes | 29,761 (40.26%) | 5,640 (53.56%) | 24,121 (38.05%) | 0.000 |
| Heart Failure | 17,842 (24.13%) | 4,019 (38.17%) | 13,823 (21.80%) | 0.000 |
| Hyperlipidemia | 45,902 (62.09%) | 8,185 (77.73%) | 37,717 (59.49%) | 0.000 |
| Hypertension | 52,834 (71.47%) | 9,474 (89.97%) | 43,360 (68.39%) | 0.000 |
| Ischemic Heart Disease | 27,793 (37.60%) | 6,001 (56.99%) | 21,792 (34.37%) | 0.000 |
| Obesity | 22,767 (30.80%) | 2,739 (26.01%) | 20,028 (31.59%) | 0.000 |
| Stroke/Transient Ischemic Attack | 20,487 (27.71%) | 4,143 (39.34%) | 16,344 (25.78%) | 0.000 |
| Tobacco Use | 16,344 (22.11%) | 1,708 (16.22%) | 14,636 (23.09%) | 0.000 |
| Traumatic Brain Injury | 5,980 (8.09%) | 720 (6.84%) | 5,260 (8.30%) | 0.000 |
| Sleep disorders | 29,691 (40.16%) | 4,323 (41.05%) | 25,368 (40.01%) | 0.045 |
| Periodontitis | 1,024 (1.39%) | 97 (0.92%) | 927 (1.46%) | 0.000 |
| Menopause | 289 (0.39%) | 13 (0.12%) | 276 (0.44%) | 0.000 |
| **MarketScan, January 2009 to June 2020, 164,148,434 patients** |  |  |  |  |
|  | **MCI** | **AD** | **MCI \ AD** | **P-value** |
| No. of patients | 424,961 (100%) | 67,973 (16.00%) | 356,988 (84.00%) | – |
| MCI age, median (IQR)<sup>c</sup> | 64 (49, 79) | 80 (73, 86) | 61 (45, 76) | 0.000 |
| Sex-female | 230,732 (54.29%) | 39,424 (58.00%) | 191,308 (53.59%) | 0.000<sup>b</sup> |
| Sex-male | 194,229 (45.71%) | 28,549 (42.00%) | 165,680 (46.41%) | – |
| Antidiabetic medication | 65,093 (15.32%) | 11,840 (17.42%) | 53,253 (14.92%) | 0.000 |
| Antihypertensives medication | 161,904 (38.10%) | 33,403 (49.14%) | 128,501 (36.00%) | 0.000 |
| Alcohol Use Disorders | 27,112 (6.38%) | 3,430 (5.05%) | 23,682 (6.63%) | 0.000 |
| Anxiety Disorders | 180,771 (42.54%) | 29,012 (42.68%) | 151,759 (42.51%) | 0.412 |
| Depression | 210,388 (49.51%) | 37,986 (55.88%) | 172,402 (48.29%) | 0.000 |
| Diabetes | 124,903 (29.39%) | 25,232 (37.12%) | 99,671 (27.92%) | 0.000 |
| Heart Failure | 81,582 (19.20%) | 21,745 (31.99%) | 59,837 (16.76%) | 0.000 |
| Hyperlipidemia | 266,840 (62.79%) | 53,061 (78.06%) | 213,779 (59.88%) | 0.000 |
| Hypertension | 282,484 (66.47%) | 59,938 (88.18%) | 222,546 (62.34%) | 0.000 |
| Ischemic Heart Disease | 135,972 (32.00%) | 33,501 (49.29%) | 102,471 (28.70%) | 0.000 |
| Obesity | 84,530 (19.89%) | 9,638 (14.18%) | 74,892 (20.98%) | 0.000 |
| Stroke/Transient Ischemic Attack | 122,531 (28.83%) | 27,784 (40.88%) | 94,747 (26.54%) | 0.000 |
| Tobacco Use | 46,010 (10.83%) | 5,145 (7.57%) | 40,865 (11.45%) | 0.000 |
| Traumatic Brain Injury | 42,635 (10.03%) | 6,255 (9.20%) | 36,380 (10.19%) | 0.000 |
| Sleep disorders | 157,272 (37.01%) | 23,566 (34.67%) | 133,706 (37.45%) | 0.000 |
| Periodontitis | 2,202 (0.52%) | 361 (0.53%) | 1,841 (0.52%) | 0.629 |
| Menopause | 4,184 (0.98%) | 328 (0.48%) | 3,856 (1.08%) | 0.000 |

<sup>a</sup> Two-sided T-test for the null hypothesis that two independent samples (population with AD diagnosis v.s. population without any AD diagnosis) have identical average values, except for sex <sup>b</sup> Chi-square test of independence of the observed male and female frequencies. <sup>c</sup> MCI age is the sample median with inter-quartile range (IQR, 25th to 75th percentile).

**Supplementary Table 2.** Selected ICD-9/10 diagnosis codes for cognitive impairment (MCI) and Alzheimer’s Disease (AD).

|  |  |
| --- | --- |
| **MCI** | **Usage:** The definition of MCI in real-world healthcare data for selection of targeted population.<br>**ICD-9 codes:**<br>331.83 Mild cognitive impairment, so stated<br>294.9 Unspecified persistent mental disorders due to conditions classified elsewhere<br>**ICD-10 codes:**<br>G31.84 Mild cognitive impairment, so stated<br>F09 Unspecified mental disorder due to known physiological condition<br>To select patients with any of above codes in database<br>**Python code:** str.startswith(('331.83', '294.9', 'G31.84', 'F09', '33183', '2949', 'G3184')) |
| **AD** | **Usage:** The definition of AD in real-world healthcare data for selection of eligible individuals before baseline and identification of outcome in follow-up.<br>**ICD-9 codes:**<br>331.0 Alzheimer's disease<br>**ICD-10 codes:**<br>G30 Alzheimer's disease<br>G30.0 Alzheimer's disease with early onset<br>G30.1 Alzheimer's disease with late onset<br>G30.8 Other Alzheimer's disease<br>G30.9 Alzheimer's disease, unspecified<br>To select patients with any of above codes in database<br>**Python code:** str.startswith(('331.0', '3310', 'G30'), na=False) |
| **AD related dementias** | **Usage:** The definition of AD related dementias in real-world healthcare data for selection of eligible individuals before baseline.<br>**ICD-9 codes:**<br>294.10 Dementia in conditions classified elsewhere without behavioral disturbance<br>294.11 Dementia in conditions classified elsewhere with behavioral disturbance<br>294.20 Dementia, unspecified, without behavioral disturbance.<br>294.21 Dementia, unspecified, with behavioral disturbance<br>290.* Dementias<br>**ICD-10 codes:**<br>F01.* Vascular dementia<br>F02.* Dementia in other diseases classified elsewhere<br>F03.* Unspecified dementia<br>To select patients with any of above codes in database<br>**Python code:** str.startswith(('F01', 'F02', 'F03', '290', '294.10', '294.11', '294.20', '294.21', '2941', '29411', '2942', '29421'), na=False) |

MCI, mild cognitive impairment; AD, Alzheimer's disease; ICD-9/10, the International Classification of Diseases 9th or 10th Revision.

**Supplementary Table 3.** Trial characteristics and estimated treatment effects ^a^ of drug candidates ^b^ from the MarketScan, 2009-2020.

| Drug | Balanced trials % | No. of treated | No. of control <sup>c</sup> | No. of unbalanced feat. | No. of unbalanced feat. after IPTW | Adjusted 2-yr survival difference % (95% CI) <sup>a</sup> | Adjusted hazard ratio (95% CI) <sup>a</sup> |
| --- | --- | --- | --- | --- | --- | --- | --- |
| gabapentin | 98 | 7625 | 15737.8 | 78.6 | 0.0 | 2.5 (2.2,2.9)*** | 0.72 (0.70,0.74)*** |
| acetaminophen | 100 | 7449 | 13282.4 | 20.9 | 0.0 | 2.4 (1.9,2.9)*** | 0.78 (0.76,0.81)*** |
| aspirin | 95 | 3412 | 10230.8 | 48.6 | 0.0 | 2.4 (2.2,2.5)*** | 0.70 (0.68,0.71)*** |
| amoxicillin | 90 | 5530 | 9996.1 | 12.3 | 0.0 | 2.0 (1.6,2.6)*** | 0.79 (0.77,0.82)*** |
| methylprednisolone | 70 | 3032 | 6976.6 | 77.6 | 0.0 | 1.8 (1.6,2.2)*** | 0.72 (0.70,0.73)*** |
| prednisone | 86 | 5557 | 10216.7 | 35.2 | 0.0 | 1.8 (1.2,2.5)*** | 0.84 (0.80,0.88)*** |
| albuterol | 89 | 4413 | 7937.2 | 20.6 | 0.0 | 1.8 (1.5,2.2)*** | 0.78 (0.76,0.80)*** |
| duloxetine | 81 | 3940 | 11820.0 | 77.0 | 0.0 | 1.6 (1.1,2.2)*** | 0.88 (0.83,0.94)*** |
| furosemide | 89 | 6067 | 11691.4 | 70.3 | 0.0 | 1.6 (1.2,2.1)*** | 0.87 (0.84,0.89)*** |
| fluticasone | 86 | 5500 | 9760.8 | 19.6 | 0.0 | 1.5 (1.0,2.1)*** | 0.86 (0.83,0.90)*** |
| bupropion | 66 | 3773 | 11319.0 | 57.8 | 0.0 | 1.4 (0.9,2.0)*** | 0.91 (0.85,0.97)* |
| triamcinolone | 95 | 2847 | 8541.0 | 17.9 | 0.0 | 1.4 (0.9,2.0)*** | 0.87 (0.85,0.89)*** |
| atorvastatin | 95 | 9161 | 18304.3 | 25.6 | 0.0 | 1.4 (1.0,1.7)*** | 0.88 (0.85,0.90)*** |
| ketoconazole | 65 | 1740 | 5220.0 | 27.8 | 0.0 | 1.3 (1.0,1.7)*** | 0.82 (0.80,0.84)*** |
| pantoprazole | 97 | 5555 | 14643.9 | 41.4 | 0.0 | 1.3 (0.9,1.8)*** | 0.92 (0.89,0.94)*** |
| zolpidem | 69 | 2617 | 5388.9 | 39.4 | 0.0 | 1.1 (0.8,1.5)*** | 0.88 (0.86,0.91)*** |
| amlodipine | 79 | 6837 | 11951.4 | 39.0 | 0.0 | 1.1 (0.6,1.6)*** | 0.93 (0.89,0.97)*** |
| tizanidine | 51 | 1648 | 4944.0 | 66.2 | 0.0 | 1.0 (0.9,1.2)*** | 0.78 (0.77,0.80)*** |
| metoprolol | 82 | 6825 | 10577.4 | 35.1 | 0.0 | 1.0 (0.5,1.6)*** | 0.95 (0.91,0.98)*** |
| omeprazole | 99 | 6966 | 14893.6 | 8.4 | 0.0 | 1.0 (0.5,1.6)*** | 0.92 (0.88,0.94)*** |
| venlafaxine | 66 | 2497 | 7491.0 | 38.2 | 0.0 | 0.9 (0.4,1.4)*** | 0.94 (0.88,1.00)<sup>ns</sup> |
| warfarin | 51 | 2764 | 3926.5 | 68.5 | 0.0 | 0.9 (0.3,1.9)** | 0.98 (0.94,1.00)<sup>ns</sup> |
| penicillin | 89 | 3808 | 8235.5 | 43.9 | 0.0 | 0.9 (0.4,1.5)*** | 0.93 (0.90,0.96)*** |
| tramadol | 96 | 5544 | 14055.0 | 37.4 | 0.0 | 0.7 (0.3,1.3)*** | 0.95 (0.92,0.98)*** |
| losartan | 87 | 5084 | 14384.5 | 34.0 | 0.0 | 0.5 (0.2,1.0)*** | 0.95 (0.92,0.97)*** |
| clotrimazole | 73 | 1651 | 4953.0 | 16.4 | 0.0 | 0.5 (0.2,0.8)*** | 0.95 (0.93,0.98)*** |
| lidocaine | 73 | 2116 | 6348.0 | 79.1 | 0.0 | 0.4 (0.2,0.7)*** | 1.00 (0.98,1.03)<sup>ns</sup> |
| mupirocin | 75 | 2165 | 6495.0 | 33.5 | 0.0 | 0.4 (0.1,0.7)*** | 0.96 (0.94,0.98)*** |
| rosuvastatin | 65 | 2663 | 7989.0 | 27.1 | 0.0 | -0.2 (-0.3,-0.1)** | 1.01 (0.99,1.02)<sup>ns</sup> |
| naproxen | 54 | 1850 | 5550.0 | 19.4 | 0.0 | -0.3 (-0.4,-0.2)*** | 1.03 (1.01,1.04)* |
| cyclobenzaprine | 61 | 2694 | 4121.7 | 34.0 | 0.0 | -0.4 (-0.5,-0.3)*** | 1.09 (1.07,1.11)*** |
| clonazepam | 69 | 2791 | 8373.0 | 64.0 | 0.0 | -0.4 (-0.5,-0.3)*** | 1.08 (1.06,1.10)*** |
| alprazolam | 88 | 2941 | 7152.6 | 22.1 | 0.0 | -0.8 (-1.1,-0.4)*** | 1.11 (1.07,1.15)*** |
| benzonatate | 58 | 2189 | 3579.0 | 50.4 | 0.0 | -0.9 (-1.1,-0.7)*** | 1.20 (1.15,1.24)*** |
| citalopram | 88 | 4298 | 12894.0 | 73.8 | 0.0 | -2.1 (-2.6,-1.5)*** | 1.29 (1.22,1.36)*** |
| mirtazapine | 87 | 3686 | 11056.1 | 49.0 | 0.0 | -2.3 (-2.8,-1.8)*** | 1.29 (1.22,1.35)*** |
| lorazepam | 93 | 3077 | 7563.9 | 33.0 | 0.0 | -2.5 (-2.9,-2.0)*** | 1.44 (1.39,1.49)*** |
| sertraline | 94 | 5524 | 16564.1 | 17.9 | 0.0 | -3.2 (-3.6,-2.8)*** | 1.47 (1.42,1.53)*** |
| escitalopram | 99 | 5041 | 15123.0 | 13.2 | 0.0 | -4.3 (-4.7,-3.8)*** | 1.55 (1.49,1.61)*** |
| quetiapine | 63 | 2934 | 5282.8 | 27.7 | 0.0 | -4.6 (-5.4,-3.9)*** | 1.58 (1.43,1.71)*** |

<sup>a</sup> 2-year standardized AD-free survival differences and hazard ratios after inverse probability of treatment re-weighting (IPTW) by regularized logistic regression-based PS model (LR-PS) using our proposed model selection strategy, adjusted for 267 covariates in total: age, sex, diagnoses codes, medications, and the time from MCI initiation date to the trial drug initiation date. Covariates were collected during baseline period. Drugs were ranked by the estimated 2-yr survival differences after IPTW. <sup>b</sup> We selected drugs with at least 50% emulated trials were balanced and for each balanced trial all the unbalanced features were balanced after IPTW. <sup>c</sup> Control groups are constructed randomly, either from alternative drug cohorts or similar drug cohorts under ATC L2. We set number of patients in the control group to maximum 3-folds as the treated group and we report the mean number of all balanced trials here. All statistics were sample means over balanced trials. Bootstrapped p-values for one-sample T-test and 1,000 bootstrapped 95% confidence interval were reported here. \*, $p < 0.05$ ; \*\*, $p < 0.01$ ; \*\*\*, $p < 0.001$ ; not significant (ns), $p \geq 0.05$ ; AD, Alzheimer's disease, MCI, mild cognitive impairment; IPTW, inverse probability of treatment re-weighting; CI, confidence interval.

**Supplementary Table 4.** Trial characteristics and estimated treatment effects ^a^ of drug candidates selected by more stringent balance criteria^b^ from the OneFlorida, 2012-2020.

| Drug | Balanced trials % | No. of treated | No. of control <sup>c</sup> | No. of unbalanced feat. | No. of unbalanced feat. after IPTW | Adjusted 2-yr survival difference % (95% CI) <sup>a</sup> | Adjusted hazard ratio (95% CI) <sup>a</sup> |
| --- | --- | --- | --- | --- | --- | --- | --- |
| gabapentin | 17 | 1237 | 3711.0 | 83.4 | 0.0 | 2.6 (2.1,3.3)*** | 0.55 (0.50,0.60)*** |
| pantoprazole | 28 | 1100 | 2840.6 | 111.1 | 0.0 | 2.1 (1.9,2.4)*** | 0.60 (0.57,0.63)*** |
| acetaminophen | 25 | 1837 | 5306.6 | 43.4 | 0.0 | 1.5 (1.2,2.0)*** | 0.74 (0.70,0.79)*** |
| atorvastatin | 38 | 1674 | 2714.1 | 71.9 | 0.0 | 1.4 (1.2,1.6)*** | 0.78 (0.75,0.81)*** |
| aspirin | 24 | 1532 | 4569.3 | 46.0 | 0.0 | -0.9 (-1.3,-0.5)** | 1.19 (1.11,1.28)** |

<sup>a</sup> 2-year standardized AD-free survival differences and hazard ratios after inverse probability of treatment re-weighting (IPTW) by regularized logistic regression-based PS model (LR-PS) using our proposed model selection strategy, adjusted for 267 covariates in total: age, sex, diagnoses codes, medications, and the time from MCI initiation date to the trial drug initiation date. Covariates were collected during baseline period. Drugs were ranked by the estimated 2-yr survival differences after IPTW. <sup>b</sup> We selected drugs with at least 10% emulated trials were balanced and we require all covariates of balanced trial should be balanced (compared with a tolerance of 2% unbalanced covariates in our primary analyses) after IPTW. <sup>c</sup> Control groups are constructed randomly, either from alternative drug cohorts or similar drug cohorts under ATC L2. We set number of patients in the control group to maximum 3-folds as the treated group and we report the mean number of all balanced trials here. All statistics were sample means over balanced trials. Bootstrapped p-values for one-sample T-test and 1,000 bootstrapped 95% confidence interval were reported here. \*, $p < 0.05$ ; \*\*, $p < 0.01$ ; \*\*\*, $p < 0.001$ ; not significant (ns), $p \geq 0.05$ ; AD, Alzheimer's disease, MCI, mild cognitive impairment; IPTW, inverse probability of treatment re-weighting; CI, confidence interval.

